# The incubation periods of Monkeypox virus clade Ib

**DOI:** 10.1101/2025.02.25.25322865

**Authors:** Javier Perez-Saez, Patrick Musole Bugeme, Megan O’Driscoll, Patrick Kazuba Bugale, Trust Faraja Mukika, Levi Bugwaja, Salomon Mashupe Shangula, Justin Bengehya, Stephanie Ngai, Antonio Isidro Carrion Martin, Jules Jackson, Noella Mulopo-Mukanya, Jackie Knee, Isabella Eckerle, Elizabeth C. Lee, Daniel Mukadi-Bamuleka, Justin Lessler, Andrew S Azman, Espoir Bwenge Malembaka

**Affiliations:** Geneva Centre for Emerging Viral Diseases, Geneva University Hospitals, Geneva, Switzerland; Department of Epidemiology, Johns Hopkins University, Baltimore, MD 21205, USA; Centre for Tropical Diseases and Global Health (CtDGH), Université Catholique de Bukavu, Bukavu, Democratic Republic of the Congo; Hôpital Général de Référence d’Uvira, Uvira Health Zone, Uvira, Democratic Republic of the Congo; Division Provinciale de la Santé Publique du Sud-Kivu, Bukavu, Democratic Republic of the Congo; Operational Centre Amsterdam (OCA), Médecins Sans Frontières, Amsterdam, the Netherlands; Médecins Sans Frontières, Uvira, DRC; Rodolphe Mérieux Institut National de Recherche Biomédicale, Goma Laboratory, Goma, Democratic Republic of the Congo; Department of Disease Control, London School of Hygiene & Tropical Medicine, London, UK; Service de Microbiologie, Département de Biologie Médicale, Cliniques Universitaires de Kinshasa, Université de Kinshasa, Kinshasa, Democratic Republic of the Congo; University of North Carolina Population Center, University of North Carolina at Chapel Hill, Chapel Hill, NC, USA; Department of Epidemiology, Gillings School of Global Public Health, University of North Carolina at Chapel Hill, Chapel Hill, NC, USA; Division of Tropical and Humanitarian Medicine, Geneva University Hospitals (HUG), Geneva, Switzerland

## Abstract

**Background:** Monkeypox virus (MPXV) clade Ib, first detected in the Democratic Republic of the Congo (DRC) in September 2023, spread internationally within months, prompting a WHO emergency declaration. Data on its incubation period, which both shapes outbreak dynamics and informs epidemic response strategies remain limited.

**Objective:** To estimate the incubation periods of mpox clade Ib examining evidence for differences by route of exposure and demographic factors.

**Design:** Bayesian analysis of clinical surveillance data collected between June and October 2024.

**Setting:** South Kivu, Democratic Republic of Congo, the epicenter of the current mpox clade Ib global outbreak.

**Participants:** Clinically attended persons with confirmed mpox clade Ib infection.

**Measurements:** Demographics, exposure history, symptom onset, and transmission route.

**Results:** Among 37 PCR-confirmed cases with high viral load (Cycle Threshold [Ct] values less than 34), the median incubation period from exposure to rash was 13.6 days (95% CrI: 10.7-20.2). Five percent of cases are expected to develop a rash within 3.8 days (95% CrI; 1.7-6.6) and 95% within 33.4 days (95% CrI: 24.1-46.4). The incubation period appeared to differ by putative transmission route: sexual transmission had a shorter median (10.3 days, 95% CrI 3.1-20.3) than non-sexual transmission (13.5 days, 95% CrI: 9.5-19.1), though the confidence intervals overlapped.

**Limitation:** Surveillance data lacked detailed exposure histories and a lower bound for exposure periods, but models accounted for these uncertainties, yielding robust median estimates.

**Conclusion:** Evidence from this study suggests that clade Ib may have a longer incubation period than other MPXV clades, and this may vary by transmission route. The shorter incubation for sexual transmission mirrors patterns seen in the predominantly sexually transmitted clade IIb outbreak, highlighting the potential role of exposure route in disease progression. These findings have implications for global recommendation on post-exposure monitoring periods and prophylaxis.

**Primary Funding Source:** This work was supported by the Gates Foundation (INV-079976) and funds from the Geneva Centre for Emerging Viruses. The funders had no role in study design, data collection and analysis, decision to publish, or preparation of the manuscript.

## Background

The Democratic Republic of Congo (DRC) is the epicenter of the current multi-country outbreak of clade I monkeypox virus (MPXV) (1). Mpox cases in South Kivu, eastern DRC, have been linked to a new MPXV subclade, clade Ib, estimated to have emerged in September 2023 (2). Early transmission chains of clade Ib were primarily attributed to sexual exposures (3). A shift to broader, non-sexual, community transmission has since been observed in parts of eastern DRC where young children are now predominantly affected (4). Community transmission of clade Ib has since been reported in 11 additional African countries (as of August 2025), including Uganda, Burundi and Kenya (5). While recent evidence has suggested fire-footed rope squirrels as a potential natural reservoir of MPXV (6) the current Clade Ib epidemic in DRC appears to be driven by transmission in humans, with little evidence of frequent spill-over events.

The differences between the natural history of clade Ib and other variants of MPXV remain unclear. The incubation period is the time between infection and symptom onset, which is linked to the speed of outbreaks and determines the feasibility and impact of control interventions (7). Estimates of the incubation period have typically ranged from 1-2 weeks though most estimates come from the predominantly sexually-transmitted clade II (8,9). Based on these estimates, the World Health Organization recommends a 21 day ‘observation period’ for risk reduction precautions and for epidemiologic studies (10). Furthermore, post-exposure vaccination is recommended by WHO for up to 14 days after exposure, which is based on estimates of the incubation period for non-clade Ib MPXV and historical data on smallpox vaccination. Changes in the incubation period of MPXV as the virus evolves may undermine these recommendations, yet we know of no estimates of the incubation period clade Ib. This gap may limit our ability to tailor efficient responses to the ongoing MPXV clade Ib epidemic.

Here, we present estimates of the incubation period of MPXV clade Ib based on case data fromSouth Kivu, DRC, focusing on its variability across demographic characteristics and putative transmission routes.

## Methods

### Study setting and population

This analysis is based on surveillance data from the Mpox Treatment Center in the Uvira health zone in South Kivu province, eastern DRC. Uvira health zone has about 460,000 inhabitants, 65% of whom reside in the city of Uvira, on the shore of Lake Tanganyika, bordering Burundi. The Uvira health zone is one of DRC’s mpox clade Ib hotspots, prioritized for interventions such as vaccination. It reported its first mpox case on May 2, 2024, eight months after the outbreak was declared in Kamituga, a remote mining town 360 km away. Initially, suspected cases, mostly adults, were managed in outpatient or community settings. However, as cases increased, an Mpox Treatment Center (MTC) was established on June 8, 2024, within the Uvira general hospital. With support from Médecins Sans Frontières, mpox care, including essential medicines and meals for patients and caregivers, became free at the Uvira MTC on June 17, 2024.

Two small-scale vaccination campaigns took place in Uvira. A two-dose vaccination campaign targeting adults perceived to be at highest risk (sex workers, health workers and contacts of cases) was conducted in October 2024 (round 1) and December 2024 (round 2), with a total of 14,000 doses of the MVA-BN vaccine available, followed by a single-dose campaign in May-June 2025 with 2,000 doses available. No other smallpox or mpox vaccination campaigns have been conducted in eastern DRC since the late 1970s (11).

### Case definitions and data collection

The official DRC Ministry of Health case definition for mpox during the time of this study included three criteria: (1) sudden high fever followed by a vesiculo-pustular rash, primarily on the face, palms, and soles; (2) the presence of at least five smallpox-type scars; or (3) fever >38.3°C, severe headache, lymphadenopathy, back pain, myalgia, and severe asthenia, followed within 1–3 days by a progressive rash spreading from the face to other body parts, including the palms and soles (12). The community case definition is broader, classifying any person with fever and a skin rash as a suspected case. At the Uvira MTC, following outbreak confirmation, a simplified case definition was applied: anyone presenting with a skin rash or papulovesicular or pustular eruptions was considered a suspected mpox case.

Starting from June 2024, the clinical staff at the Uvira General Hospital’s Mpox Treatment Centre (MTC) conducted structured interviews with suspected cases. Data were collected on demographics, exposure history, and clinical manifestations, using forms on the KoboCollect platform. A suspected case contact was defined as someone with a rash, lesions and/or similar symptoms to the patient or a confirmed mpox case, contacted in the three weeks prior to symptom onset or diagnosis of the clinical case. For each reported contact with a suspected case, we collected information on whether a single or multiple exposures occurred, the type(s) of exposure (physical non-sexual contact, sexual contact, respiratory contact, contact with contaminated material; see Supplementary Material Section S1), and the date of most recent exposure.

MTC staff collected skin lesion secretions of suspected cases using a dry swab, whereas oropharyngeal secretions were taken with a swab that was soaked into a viral transport medium. While lesion swabs were meant to be collected and tested from all suspected cases, only a subset had test results due to stockouts of materials and reagents during this phase of the outbreak. The subset of suspected cases that were tested had dry lesion swabs tested by qPCR with GeneXpert (Xpert® Mpox; Cepheid, Sunnyvale, USA) or the Radi^®^ Fast Mpox Kits (KH Medical, Pyeongtaek-si, Gyeonggido, Republic of Korea). The choice of PCR test was based on the availability of reagents and lab testing capacity during the study period and was not based on patient characteristics.

Based on previous reports of potential misclassification of patients with high PCR cycle threshold (Ct) values, we defined a subset of cases who had Ct values below a conservative threshold for positivity (<34) to reduce the risk of false positives (13). We refer to these individuals as “high-confidence confirmed cases,” those with a Ct value below 39 as “confirmed cases”, and all clinical cases as “suspected cases”.

Due to the deletion of the OPG032 gene in clade Ib viruses, amplification of the Genexpert MPXV PCR target is inhibited (2). Clade Ib viruses therefore test negative for MPXV by Genexpert assay but positive for OPXV (based on the E9L target). This result is specific to Clade Ib viruses and was seen for all Genexpert tests conducted in Uvira. In addition, a subsample of 49 cases from Uvira from the same period, were clade-typed using the TIB Molbiol® qPCR kit (TIB Molbiol, Eresburgstrasse, Berlin, Germany), confirmed that all cases tested belonged to clade Ib (14,15).

### Statistical inference of the incubation period

The aim of the statistical modeling framework was to infer the distribution of the incubation periods of MPXV clade Ib for different definitions of symptom onset (fever, rash, earliest symptom), and its variability across key factors, including demographic characteristics and transmission routes. To do so, we used Bayesian methods to obtain a robust estimate of the incubation period distribution accounting for the limitations of the data and known sources of bias in the estimation of epidemiologic delay distributions (16).

Our modeling framework accounted for multiple reported contacts per case, censored times from exposure to symptom onset as only time to most recent contact was reported, and right-truncation of the time from exposure to symptom onset as only contacts in the previous 21 days were recorded). When case contacts were also enrolled in our study, we used the date of rash onset of contacts to bound the exposure window, and assumed a uniform probability of exposure within that window. When the exposure time was unknown and contact pair information unavailable, we used a maximum exposure window of 35 days prior to symptom onset up to the time of most recent contact, consistent with published estimates of the incubation period for clades I and II (8). We used published estimates of the time to clade Ib lesion resolution (17) to set priors on the date of exposure within possible exposure windows. We compared across multiple distributional assumptions typically used for incubation period estimation (Log-normal, Gamma and Weibull), and across different assumptions on the probability of unreported exposures in the community based on the observed epidemic curve (no additional community exposures vs. exposures as a function of the epidemic curve). We set priors on the mean incubation period based on published estimates of the clade IIb 2022 epidemic and historical clade I and clade II outbreaks in Ponce et al. (2024) (8). A full model description is given in Supplementary Methods Section S2.

We produced separate incubation period estimates for different symptom types: fever, rash, and earliest symptom. In addition, we produced stratified estimates by demographic characteristics (age group, sex), exposure type (sexual vs. non-sexual, see definitions in Supplementary Material Section S1.1), epidemic period (before vs. after to Sep 1st, 2024, Supplementary Figure S2), and hospitalization status (inpatient vs. outpatient). As multiple exposure types could be reported for the same contact, we defined the exposure to be “sexual” if any of the exposure types was sexual for that contact, and “non-sexual” otherwise. Since all cases were not tested and current diagnostics are imperfect, we conducted all analyses using three sets of outcomes: (i) all suspected cases, (ii) all cases with positive PCR result (“confirmed cases”) and (iii) all confirmed cases with a Ct value less than 34 (“high-confidence confirmed cases”) (see previous Section “Case definitions and data collection”).

Inference was drawn using a Hamiltonian Monte Carlo sampler as implemented in the Stanprogramming language. Model comparison was performed using estimated leave-one-out crossvalidation log-predictive density (18).

## Results

### Clinical surveillance and reported exposures of mpox cases

Of the 973 cases recorded between June and October 2024, 35.6% (346/973) of suspected mpox cases reported having contact with a suspected mpox case, with 70% (243/346) of them reporting temporal information of these contacts. Of the 243 suspected cases with reported contacts, 48% (117/243) were male, 30% (73/243) were children under 5, and 2% (5/243) were 45 and older, thus susceptible to having received smallpox vaccine through routine vaccination (11) (Table 1). Only 1% (3/243) reported having a smallpox vaccine. The dates of vaccination were not reported but all 3 individuals were older than 48 years, therefore could have been vaccinated as children during smallpox eradication efforts. Two had a PCR result, both were positive and one had a Ct value less than 34. A total of 44% (107/243) of cases with reported contacts were hospitalized, and 3% (8/243) were tested for HIV, all of which were negative with one on pre-exposure prophylaxis. Among the cases with both complete contact information and valid laboratory results (123/243), 72% (92/123) were PCR-positive (“confirmed cases”) and 25% (31/123) were negative. Among confirmed - cases 40% (37/92) were considered high-confidence confirmed cases based on their low Ct values, “high confidence confirmed cases” (13). The distribution of cases by age, sex and self-reported exposure types were similar across these three case definitions (Table 1).

**Table 1.**
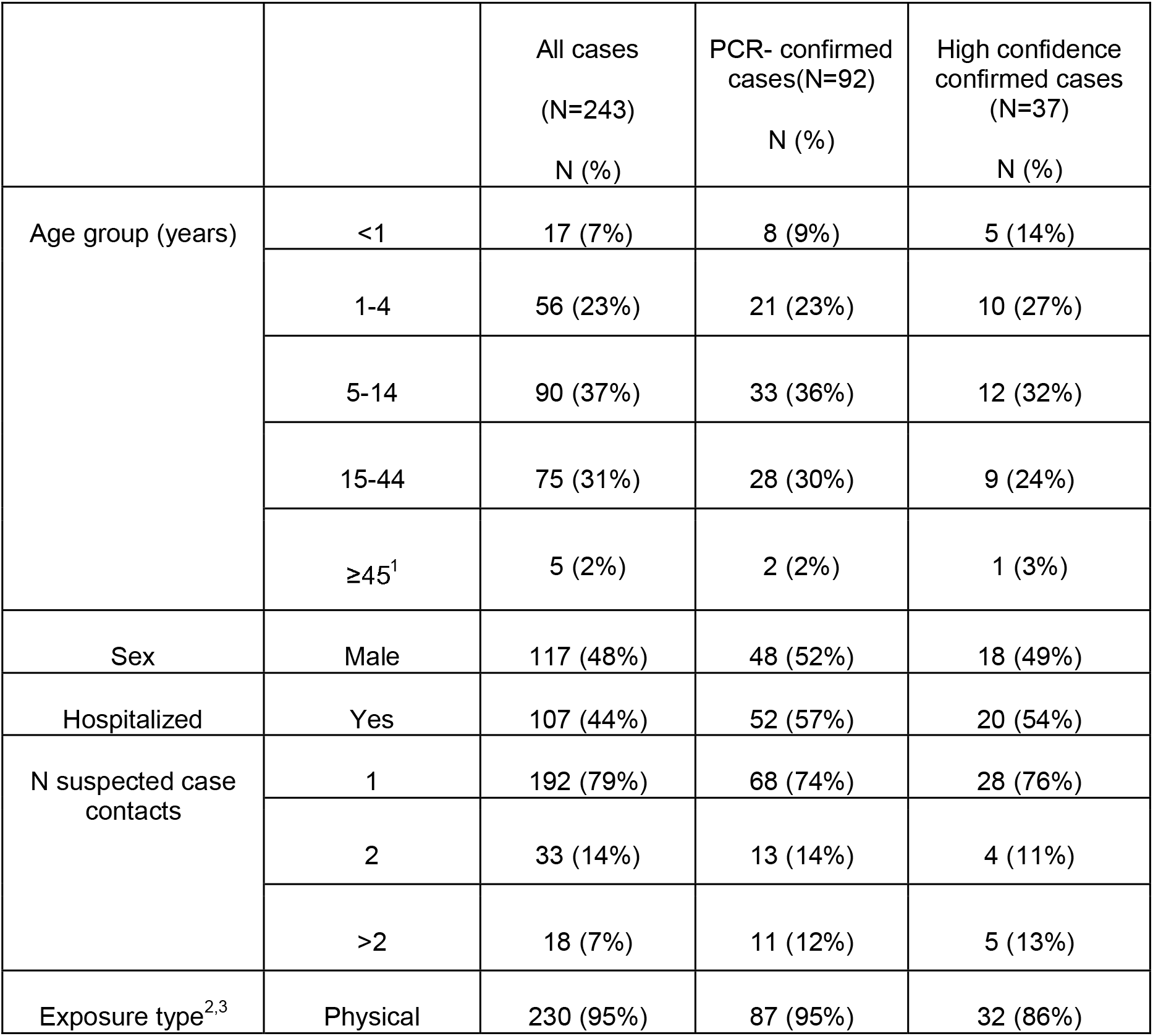
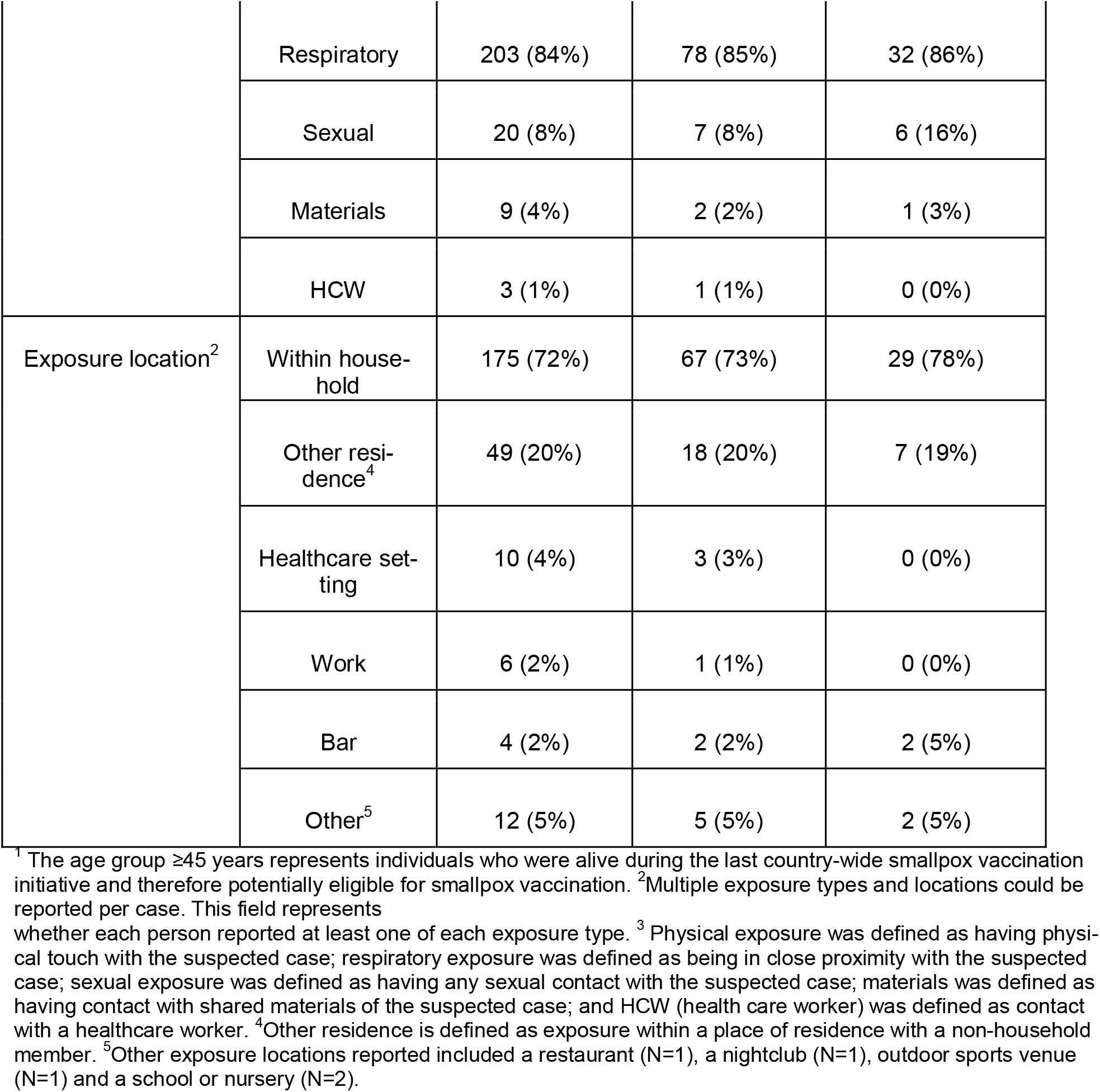
Mpox patient demographics and exposure details for all cases (N=243), PCR-confirmed cases (N=92) and high confidence (Ct value <34) (N=37).

|  |  | All cases<br>(N=243)<br>N (%) | PCR- confirmed<br>cases(N=92)<br>N (%) | High confidence<br>confirmed cases<br>(N=37)<br>N (%) |
| --- | --- | --- | --- | --- |
| Age group (years) | <1 | 17 (7%) | 8 (9%) | 5 (14%) |
|  | 1-4 | 56 (23%) | 21 (23%) | 10 (27%) |
|  | 5-14 | 90 (37%) | 33 (36%) | 12 (32%) |
|  | 15-44 | 75 (31%) | 28 (30%) | 9 (24%) |
|  | ≥45<sup>1</sup> | 5 (2%) | 2 (2%) | 1 (3%) |
| Sex | Male | 117 (48%) | 48 (52%) | 18 (49%) |
| Hospitalized | Yes | 107 (44%) | 52 (57%) | 20 (54%) |
| N suspected case<br>contacts | 1 | 192 (79%) | 68 (74%) | 28 (76%) |
|  | 2 | 33 (14%) | 13 (14%) | 4 (11%) |
|  | >2 | 18 (7%) | 11 (12%) | 5 (13%) |
| Exposure type<sup>2,3</sup> | Physical | 230 (95%) | 87 (95%) | 32 (86%) |
|  | Respiratory | 203 (84%) | 78 (85%) | 32 (86%) |
|  | Sexual | 20 (8%) | 7 (8%) | 6 (16%) |
|  | Materials | 9 (4%) | 2 (2%) | 1 (3%) |
|  | HCW | 3 (1%) | 1 (1%) | 0 (0%) |
| Exposure location<sup>2</sup> | Within house-<br>hold | 175 (72%) | 67 (73%) | 29 (78%) |
|  | Other resi-<br>dence<sup>4</sup> | 49 (20%) | 18 (20%) | 7 (19%) |
|  | Healthcare set-<br>ting | 10 (4%) | 3 (3%) | 0 (0%) |
|  | Work | 6 (2%) | 1 (1%) | 0 (0%) |
|  | Bar | 4 (2%) | 2 (2%) | 2 (5%) |
|  | Other<sup>5</sup> | 12 (5%) | 5 (5%) | 2 (5%) |

<sup>1</sup> The age group ≥45 years represents individuals who were alive during the last country-wide smallpox vaccination initiative and therefore potentially eligible for smallpox vaccination. <sup>2</sup>Multiple exposure types and locations could be reported per case. This field represents whether each person reported at least one of each exposure type. <sup>3</sup> Physical exposure was defined as having physical touch with the suspected case; respiratory exposure was defined as being in close proximity with the suspected case; sexual exposure was defined as having any sexual contact with the suspected case; materials was defined as having contact with shared materials of the suspected case; and HCW (health care worker) was defined as contact with a healthcare worker. <sup>4</sup>Other residence is defined as exposure within a place of residence with a non-household member. <sup>5</sup>Other exposure locations reported included a restaurant (N=1), a nightclub (N=1), outdoor sports venue (N=1) and a school or nursery (N=2).

The 243 suspected cases with complete contact information reported contact with 320 suspected mpox cases. 79% (192/243) reported contact with a single suspected case and 21% (51/243) to more than one suspected case (Table 1). The majority of suspected cases reported non-sexual physical contact with a case (95%, 230/243) and/or respiratory contact (84%, 203/243), with most exposures reported to have occurred within the household. Only 8% (20/243) of cases reported sexual exposures. Exposure types among suspected and confirmed cases was similar, though sexual exposure was more frequent among high-confidence confirmed cases (16% vs 8% in suspected and confirmed cases, shown in Table 1).

### Incubation period estimation

We estimated the incubation period based on data from 92 confirmed cases with complete data on reported exposure to suspected cases, with a focus on 37 high confidence confirmed cases (see Methods). We performed secondary analyses on all 243 suspected cases with contact information as not all cases were tested and the field sensitivity of current PCR assays is not well understood. Among confirmed cases cases, the median time from most recent contact (before symptoms) to rash onset was 10 days (Interquartile Range, IQR: 3–14), 9 days (IQR: 3–14) to fever onset, and 9 days (IQR: 2–13) to any symptoms, with similar times for high confidence and suspected cases (Supplementary Figure S3). However, these numbers do not account for biases due the fact that the last reported exposure may not have been the one that led to infection.

When restricting analyses to high confidence confirmed cases, we estimate that the median time from exposure to rash onset is 13.6 days (95% CrI: 9.6-19.0) (Figure 1, Table 2), with 5% of infections expected to develop rash within 3.1 days of exposure (95% CrI; 1.3-5.5), and 95% within 32.3 days (95% CrI: 22.4-45.8). Estimates of the median time to onset of fever were similar (14.5 days; 95% CrI: 9.7-20.5) and shorter for the onset of any symptom (11.6 days; 95% CrI: 7.7-16.3) (Table 2). Estimates based on alternative models, including those with different distributional assumptions, were like those with the best fitting model (Supplementary Tables S1-S2).

**Table 2:** Estimated quantiles of incubation period distribution by symptom type.

|  |  |  | Incubation period quantile in days (mean, 95% CrI) |  |  |  |  |
| --- | --- | --- | --- | --- | --- | --- | --- |
| PCR confirmation | Symptom type | N | 5% | 25% | 50% | 75% | 95% |
| High confidence cases | any symptom | 37 | 1.9 (0.7-3.8) | 6.2 (3.4-9.7) | 11.6 (7.7-16.3) | 19.0 (13.6-25.8) | 33.2 (22.9-46.5) |
|  | fever | 28 | 3.8 (1.3-7.2) | 9.1 (5.1-14.0) | 14.5 (9.7-20.5) | 21.0 (15.4-28.8) | 31.9 (22.7-45.0) |
|  | rash | 37 | 3.1 (1.3-5.5) | 8.2 (5.0-12.1) | 13.6 (9.6-19.0) | 20.5 (15.0-28.1) | 32.3 (22.4-45.8) |
| Confirmed cases | any symptom | 92 | 3.2 (1.8-4.9) | 8.9 (6.5-11.8) | 15.1 (12.0-19.0) | 23.0 (18.6-28.7) | 36.8 (28.7-47.5) |
|  | fever | 77 | 3.4 (2.0-5.2) | 9.1 (6.6-12.1) | 15.3 (12.0-19.5) | 22.9 (18.4-29.2) | 36.1 (27.9-47.1) |
|  | rash | 92 | 4.8 (3.0-6.8) | 11.3 (8.7-14.4) | 17.6 (14.3-21.8) | 25.0 (20.5-31.1) | 37.1 (29.6-47.0) |
| Suspected cases | any symptom | 243 | 2.9 (2.0-3.8) | 8.6 (7.1-10.3) | 15.2 (12.9-17.9) | 23.7 (20.3-28.1) | 38.9 (32.5-47.7) |
|  | fever | 195 | 3.9 (2.8-5.1) | 9.9 (8.1-11.9) | 16.0 (13.7-18.8) | 23.4 (20.0-27.8) | 35.7 (29.6-44.0) |
|  | rash | 243 | 4.1 (3.1-5.3) | 10.5 (8.9-12.4) | 17.0 (14.8-19.7) | 24.9 (21.6-29.1) | 38.1 (32.1-45.8) |

Main model estimates (Weibull distribution, without community infections), shown for PCR-positive cases (main results, n=92) and all patients with reported contacts (n=243) by symptom type. Estimates for all distributional assumptions are provided in the Supplementary Material. High confidence positives were defined as those with a Ct value lower than 34.

**Figure 1.**
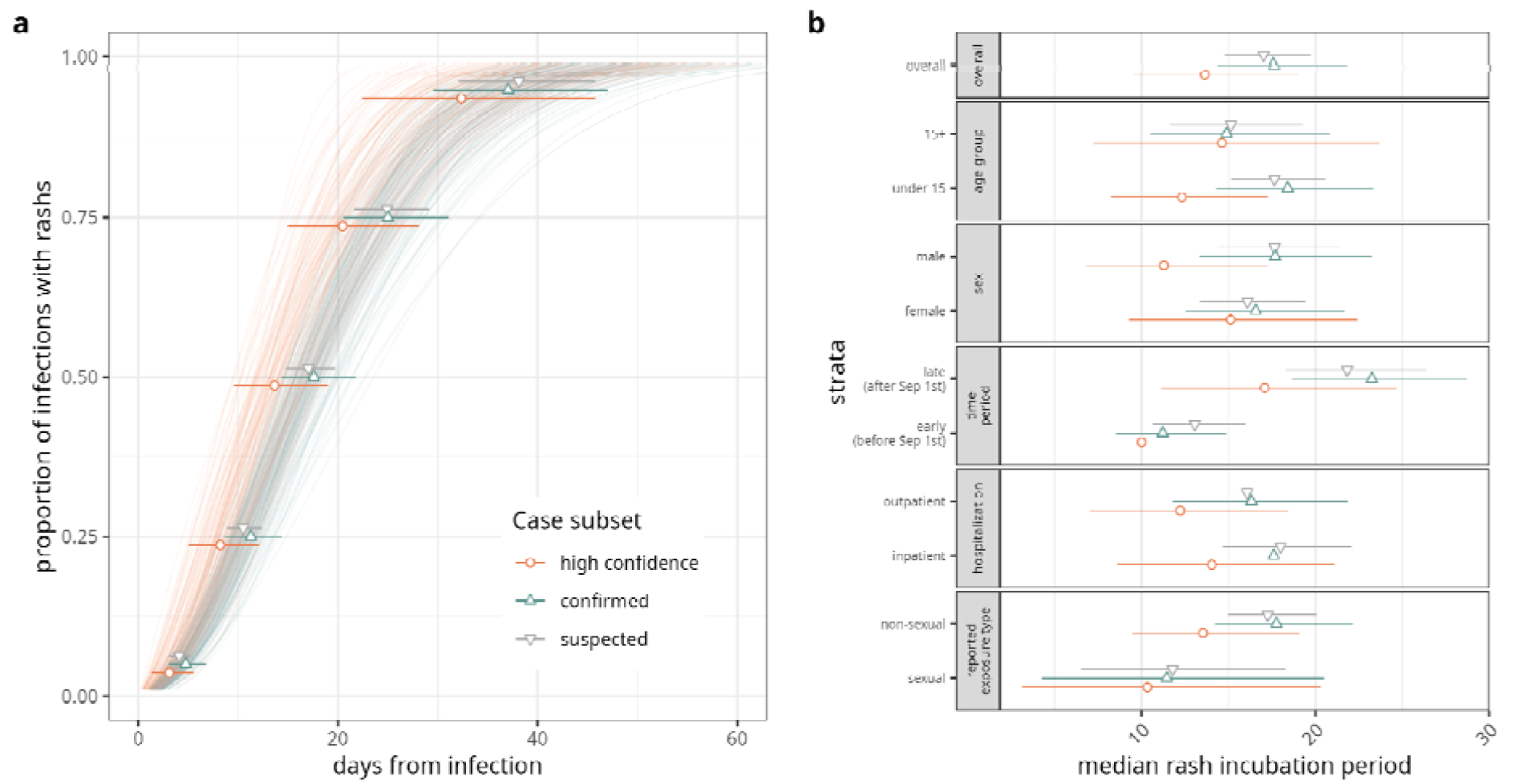
Estimates of Mpox clade Ib rash incubation period. a) Posterior draws of the incubation period cumulative distribution function (lines, 100 draws) and estimates of quantiles of interest (points: mean, bars: 95% credible intervals, CrI) by mpox case definition. Estimates are shown for the Weibull distribution assumption, and the main model (no community infections), which was retained in model comparison. b) Estimates of the median incubation period (dots: mean, bars: 95% CrI) by strata for the overall estimates (first facet) and across analysis stratifications.

We found evidence of differences in the incubation period by transmission route (Figure 1b, Table 3). The median rash incubation period was shorter for putative sexual vs. non-sexual transmission (median 10.3 days, 95% CrI: 3.1-20.3, vs. 13.5, 95% CrI: 9.5-19.1) (Table 3, Supplementary Material Section S1.2), although the 95% credible intervals overlapped. We estimate a shorter rash incubation period in the early phase of the epidemic (prior to Sep 1st, median 10.0 days, 95% CrI: 5.7-15.8) versus the later phase (after Sep 1st, median 17.1 days, 95 % CrI: 11.1-24.7). Incubation periods among those less than 15 years old appeared shorter that those 15 years and above (10.0 days, 95% Cr: 6.0-15.4, vs. 13.7, 95% CrI: 6.7-22.3; Supplementary Tables S3-S4), though this trend reversed when considering alternative case definitions. We found similar differences between groups in stratified estimates for time to fever and any symptom onset (Figure S4, Supplementary Tables S3-S5).

**Table 3:**
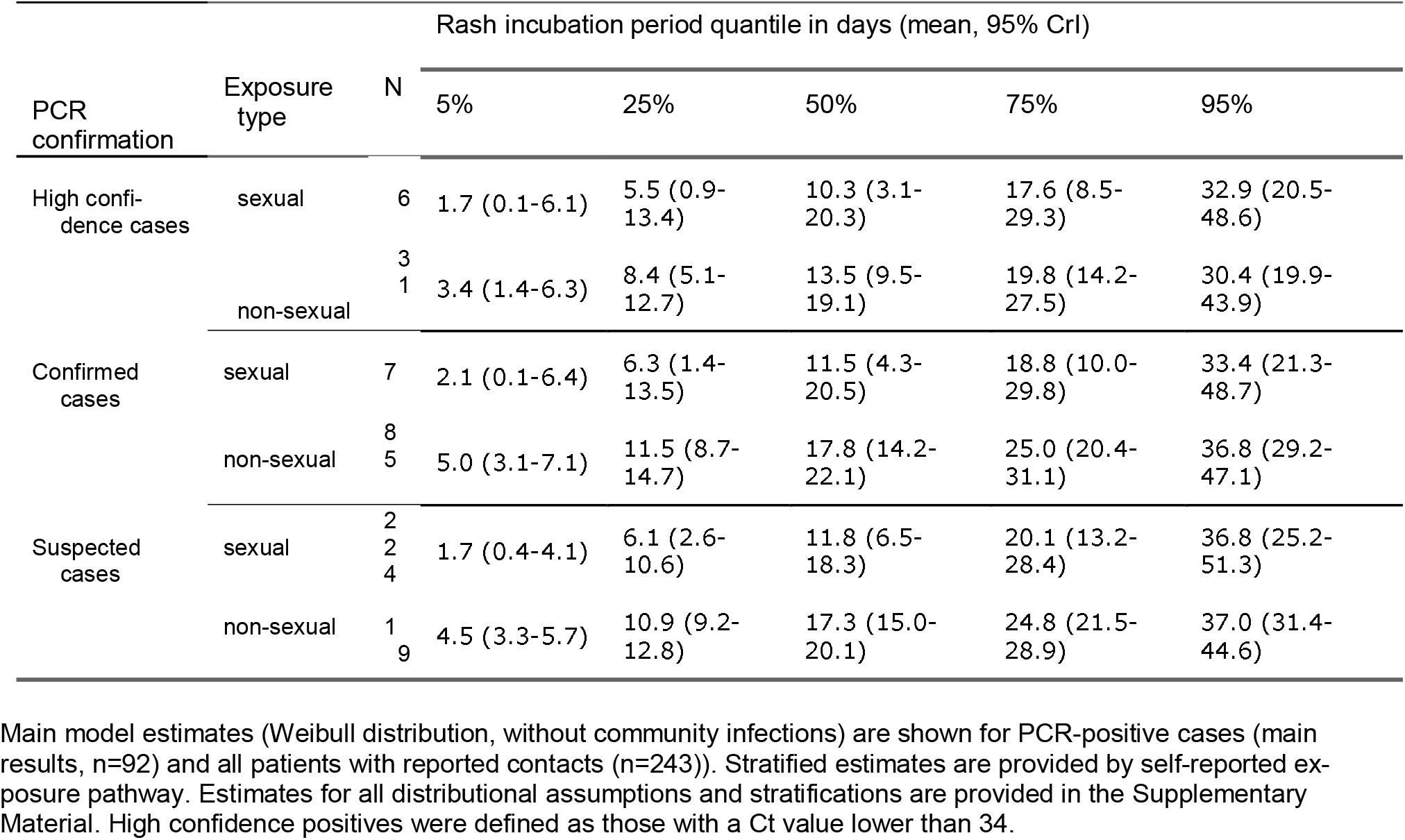
Estimated quantiles of incubation period distribution to rash onset by exposure type.

### Sensitivity Analyses

Estimates of the median incubation period using confirmed (17.6 days; 95% CrI: 14.3-21.8) and suspected cases (17.0 days; 95% CrI: 14.8-19.7) were longer than our main estimates from high-confidence confirmed cases. The median incubation period was similar for self-reported sexual exposure across case definitions (confirmed: 11.5, 95% CrI: 4.3-20.5; suspected: 11.8, 95% CrI: 6.5-18.3) but was consistently longer for non-sexual exposure (confirmed: 17.8, 95% CrI: 14.2-22.1; suspected: 17.3, 95% CrI: 15.0-20.1, Supplementary Table S3-S5). Our median incubation period estimates for high-confidence confirmed cases was robust to the choice of Ct thresholds when these were smaller than or equal to 34 (used for our main results) but increased with increasing Ct thresholds above 34 (Supplementary Figure S5).

We further refined our analysis to 50 contact pairs for which symptom onset dates were available for both case and contact, all-but-one of which reported non-sexual exposures (Supplementary Table S8). Median incubation period estimates based on all contact pairs were similar to our main estimates for non-sexual exposure (N=50, median 12.4 days, 95% CrI: 9.2-16.5), but shorter for high-confidence cases (N=7, median 9.8 days, 95% CrI: 2.7-20.3) and slightly longer for confirmed cases (N=19, median 15.5 days, 95% CrI: 9.3-23.1), although these case definitions had limited sample sizes (Supplementary Figure S6, Supplementary Table S9). We were unable to perform stratified analyses on exposure type within this subset to the limited sample size of reported sexual exposures.

## Discussion

We estimate the median incubation period for MPXV clade Ib in Uvira, DRC to be 13.5 days. This is longer than estimates for MPXV clades Ia or IIb (9–10 days) (19), and slightly longer than, but not inconsistent with, historical estimates of the incubation period of smallpox (20). We find evidence of differential incubation period by route of transmission.

The incubation period from sexual contact was lower (10.5 days) than for non-sexual exposure (13.5 days), although the uncertainty intervals overlapped. Our estimate of clade Ib incubation periods align with those in a Clade Ib outbreak in Kamituga, another region of the DRC, where transmission was predominantly heterosexual and in adults (21), and were similar to estimates for the predominantly sexually transmitted MPXV clade IIb 2022 outbreak (8) (Supplementary Figure S6). By contrast, our median incubation period estimates from non-sexual exposures are longer than the reported range of 5-13 days for non-sexual human-to-human transmission events during a clade Ia outbreak in 2013 in central DRC (9), but are compatible with earlier studies reporting clade Ia non-sexual human-to-human incubation periods in the country, ranging between one and three weeks (22,23). Comparison with Clade IIa zoonotic exposures in the 2003 U.S. outbreak (median 12 days, range 1–41, mostly adults) further illustrates that both host age and exposure route are important determinants of incubation period (24).

The fact that previous MPXV clade IIb incubation period estimates align closely with our clade Ib results for sexual contacts, but less with the longer overall estimates, may be at least in part attributable to biological factors. Higher infectious doses are known to shorten the incubation periods of viral infections (25) and route of exposure (mucosal vs. epidermal) may also impact rash onset timing as mpox lesions preferentially appear at anatomical sites correlating with site of exposure (26). Taken together, our exposure-stratified clade Ib results and previous clade IIb estimates thus support the hypothesis that MPXV incubation periods are shorter after sexual exposures due to higher viral infectious doses and differential exposure routes than for non-sexual contacts.

In Uvira, we find that the mode of transmission appears to have changed over the course of the epidemic, from predominantly sexual in the early weeks to mainly non-sexual in the later (4). These shifts in transmission may therefore explain our longer overall incubation period estimates, as most cases for which we had contact information were recruited in the later phase of the epidemic. Our longer estimates may also stem from methodological differences, as we here correct for common sources of downward-bias (16), including censoring due to unknown exact contact dates, and right-truncation as we only asked about exposures in the past 21 days following WHO recommendations (27).

Our incubation period estimates were sensitive to PCR-based case definitions, which may have implications for MPXV clade Ib epidemiologic studies and control. The fact that suspected and confirmed cases had similarly longer incubation periods with respect to our high-confidence confirmation definition may indicate low test specificity when using higher Ct value thresholds to define positivity. These findings echo challenges in defining positive cases during the clade IIb outbreak in the USA (13). Taken together, these elements warrant careful investigations of Ct value distributions in suspected MPXV clade Ib cases and their implication for defining positivity thresholds.

Our results have implications for both surveillance and control of mpox clade Ib. Around one-fifth of cases (23.2%, 95% CrI: 7.2-43.6, Supplementary Table S5) developed rash after the WHO’s recommended three-week monitoring period for exposed individuals and recall period for epidemiologic investigations. As detailed below, this finding is subject to limitations in our study and is to be interpreted with care, especially given the wide uncertainty intervals. Nevertheless, these results suggest that locally adapted time bounds could improve our ability to both understand and ultimately contain this virus. Additionally, our observation that the median incubation period may be longer than previous estimates for MPXV, suggests the possibility that the time-window for post-exposure prophylactic vaccination could be longer than current recommendations, though evidence on the effectiveness of this use of the vaccine is needed.

These results come with several limitations. First, we attribute these estimates to clade Ib, yet cases were not all clade typed. However, all 45 samples from Uvira that were clade typed with Tibmolbiol^®^ qPCR clade typing kits (TIB Molbiol, Eresburgstrasse, Berlin, Germany) were clade Ib, as were all samples in the province of South Kivu that were clade typed at INRB Goma. Second, data were collected during routine surveillance activities, allowing for limited descriptions of contacts and exposures. We only asked about the date of most recent exposure to a suspected mpox case and do not have the lower bound on the exposure period. We accounted for these sources of uncertainty in the models and results were robust to various assumptions about this lower bound. Third, our analyses implicitly assumes that transmission is from person-to-person, with no zoonotic infections. We have no data suggesting a major role for zoonotic infections in this clade Ib outbreak, and it is unclear how these might influence our estimates of the incubation period. Finally, our estimates of the right tails of the incubation period have more uncertainty than the median because of their sensitivity to distributional assumptions.

Our results illustrate that the incubation period of MPXV clade Ib may be longer than previously characterized MPXV variants, and may vary by transmission route, which has important implications for outbreak management. The observed shorter incubation periods for sexual vs. non-sexual exposures suggests that dose and exposure site may have a major influence on the timeline of disease, underscoring the need for flexible and locally adapted public health responses. Our estimates suggest that the current three-week monitoring period recommended by WHO may be insufficient for a non-negligible fraction of clade Ib non-sexually transmitted infections, potentially leading to missed cases and ongoing transmission, although this finding should be confirmed by future MPXV clade Ib studies. Strengthened surveillance, extended monitoring periods, and targeted interventions based on dominant transmission routes could improve containment efforts in South Kivu, DRC, and other clade Ib-affected countries. Future research should aim to confirm these findings in other regions and further explore the ways in which viral dose and exposure site may influence the natural history of disease.

## Data Availability

All data produced in the present study are available upon reasonable request to the authors.

## Patient Consent Information

The data used in this study were collected with the Uvira health zone team for public health surveillance. Ethical approvals were obtained from the Institutional Review Boards of the Johns Hopkins Bloomberg School of Public Health (reference number IRB00030442) and the Université Catholique de Bukavu (UCB/CIES/NC/019/2024) to use those data.

## Author Contributions

Conceptualization: ASA, EBM; Data curation: PMB, JJ, EBM; Formal Analysis: JP; Funding acquisition: SN, IC, ASA, EBM; Investigation: PMB, PKB, TFM, LB, SMS, NM; Methodology: JP; Project administration: PMB, LB, ASA, EBM; Resources: JB, IE, DM, EBM; Software: JP, MO, DM; Supervision: PMB, LB, JB, ASA, EBM; Validation: JB, DM, EBM; Visualization:; Writing – original draft: JP, MO, ASA, EBM; Writing – review & editing: JP, PMB, PKB, TFM, LB, SMS, JB, SN, IC, JJ, NM, JK, IE, ECL, DM, JL, ASA, EBM

