## Supplementary file for "The incubation periods of Monkeypox virus clade Ib"

### S1 Study description

#### S1.1 Contact information and exposure definition

Detailed contact information was only available for a subset of cases where the in-depth clinical surveillance form had been administered.

A suspected contact was defined as anyone who had similar symptoms or was a probable or confirmed mpox case with whom the patient had contact within three weeks before symptom onset or diagnosis. For patients reporting exposure to suspected cases, we documented:

- The number of suspected contacts they had been in contact with.
- Whether the contact was repeated or a single occurrence
- The estimated duration of contact
- The relationship between the patient and the reported contact. A sexual relationship was defined as contact with a reported spouse or sexual partner.

In addition to documenting relationships between patients and their contacts, we collected data on the place and type of contact. Contact types were classified as follows:

- **Physical non-sexual contact:** Skin-to-skin or skin-to-mucosal contact, including touching, hugging, or kissing on the cheek.
- **Sexual contact:** Mucosal contact, including kissing on the mouth, as well as oral, vaginal, or anal sexual intercourse. In this analysis, sexual contact was only considered for individuals aged 15 years or older.
- **Respiratory contact:** Prolonged face-to-face exposure to a case in proximity for at least 15 minutes, including activities such as eating together, engaging in close frontal conversations, sitting next to an infected person while talking, or sharing a vehicle.
- **Contact with contaminated materials:** Exposure to fluids or dislodged lesion material (crusts) through the skin or mucous membranes, such as handling contaminated clothing or bedding without PPE, cleaning infected rooms without PPE, sustaining a needlestick injury from a case sample, or handling a case sample during laboratory activities.

### S2 Modeling framework

Code is available at https://github.com/HopkinsIDD/mpox_cladeIb_incubation_period.

#### S2.1 Model v0: Base model

##### S2.1.1 Single conatct

The base model infers the incubation period accounting for right-censoring of the time to exposure. For some patients we know that there was a single exposure with the symptomatic contact. In this case the observation likelihood of parameter vector $\theta$ parametrizing the pdf of the incubation period, $f_I(\tau)$, where $\tau$ is the time from infection to symptom onset, is:

$$L_i(\theta) = f_I(t_{S_i} - t_{E_i}|\theta),$$

where $t_{S_i}$ is the time of symptom onset of patient $i$, and $t_{E_i}$ is the time of single exposure to it's symptomatic contact. As a first pass we take $f_I$ to be a lognormal distribution with mean $\mu$ and sd $\sigma$.

#### S2.2 Censoring

Most patients however report multiple exposures per contact, and we only have the date of most recent contact. This means that the time from exposure to symptom onset is right-censored (we only know that the incubation period is at least a given value). Assuming a plausible lower (left) bound for the time of exposure

$t_{E_l,i}$, the reported time of last exposure corresponds to the upper (right) bound, $t_{E_r,i}$, and the likelihood can be written as:

$$L_i(\theta) = \int_{t_{E_{l,i}}}^{t_{E_{r,i}}} \phi(t) f_I(t_{S_i} - t|\theta) dt, \quad (1)$$

where $\phi(t)$ is the probability that the infectious exposure occurred at time $t_{E_{l,i}} \leq t \leq t_{E_{r,i}}$.

When contact pair information was available within our dataset (see section S3), we used the date of rash onset of the contact as a lower bound of the exposure window, and assumed a uniform probability of exposure during the exposure window.

In the absence of contact pair information, we here assume that the probability of a given exposure time is proportional to the probability that the infector contact had already developed lesions at time $t$:

$$\phi(t) = \frac{1 - F_L(t_{E_{r,i}} - t)}{\int_{t_{E_{l,i}}}^{t_{E_{r,i}}} 1 - F_L(t_{E_{r,i}} - \tau) d\tau},$$

where $F_L(t)$ is the cumulative distribution function of time to lesion resolution. We fit a lognormal distribution to the survival curve reported in Brosius et al. [1]. The optimal parameters are a logmean of log(14.1) days and logsd of 0.5 days.

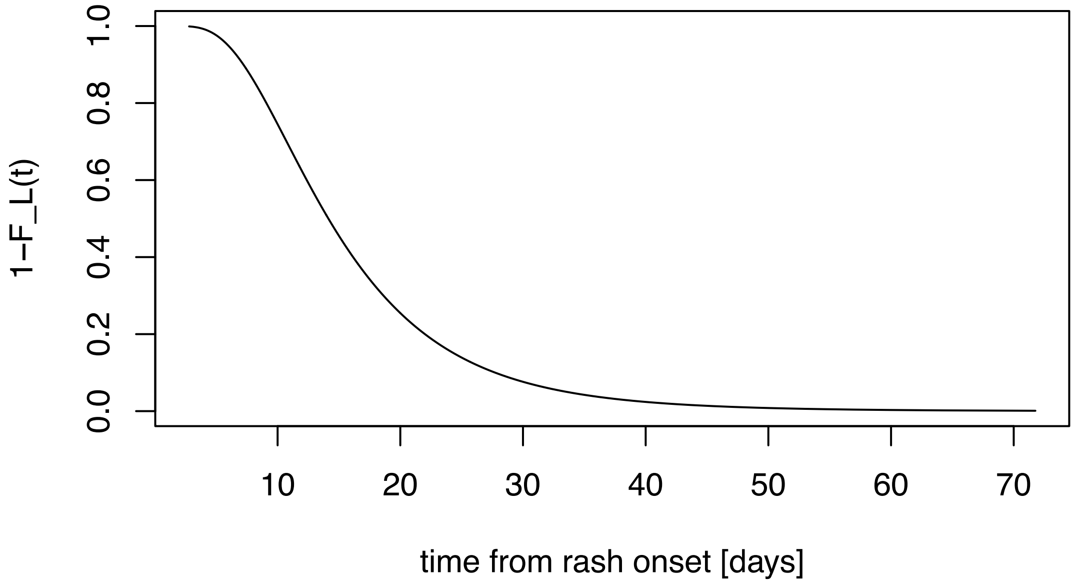

Figure S1: CCDF of time to lesion resolution

Based on this estimate, we bound possible exposure times to 35 prior to most recent exposure, $t_{E_{l,i}} = t_{E_{r,i}} - 35$, consistent with previous estimates of the incubation period of MPXV Ponce et al. [2]. This corresponds to a 90% probability of having resolved symptoms the time of exposure.

##### S2.2.1 Truncation

Study participants were asked to report contacts in the three weeks prior to symptom onset, following WHO recommendations. As such, it is necessary to account for right-truncation of the incubation period, in addition to censoring. Following Park et al. [3] the likelihood of a truncated observation, $L_i^T(\theta)$, is:

$$L_i^T(\theta) = \frac{\int_{t_{E_{l,i}}}^{t_{E_{r,i}}} \phi(t) f_I(t_{S_i} - t|\theta)dt}{\int_{t_{E_{l,i}}}^{t_{E_{r,i}}} \phi(t) F_I(t_{E_{r,i}} + 21 - t|\theta)dt},$$

where $F_I$ is the cumulative distribution function of the incubation period. In our implementation we also account for daily censoring arising from the reporting of delays at discrete daily time intervals Park et al. [3].

Although the questionnaire only asked about contacts in the 21 days prior to symptom onset, 28 patients of 973 with contact information reported exposure that occurred more than 21 days prior to symptom onset. This indicates that patients may not of taken the 21-day cutoff into account when answering the questionnaire, which means that truncation may not be justified for all contacts.

As the true probability of non-adherence to the 21-day cutoff is unknown, we jointly inferred it in our Bayesian modeling framework. We denote the underlying cutoff non-adherence probability as $\xi$, $L_i^U(\theta)$ the un-truncated likelihood in eq. (1), and $L_i^T(\theta)$ the truncated likelihood of observation $i$. Marginalizing over cutoff non-adherence, we have the the final likelihood is:

$$L_i(\theta) = (1 - \xi) \times L_i^U(\theta) + \xi \times L_i^T(\theta). \quad (2)$$

To inform the cutoff non-adherence probability, we link it to the number of raw incubation period observations longer than 21 days as:

$$L(\xi) = Binomial\_CCDF(n_{21+}|N, \xi),$$

where $Binomial\_CCDF$ is the complementary cumulative distribution function of the binomial distribution, $n_{21+}$ is the number of reported contacts more than 21 days prior to symptom onset, and $N$ is the total number of reported contacts.

##### S2.2.2 Mulitple conatcts

Around 7% of all patients (70/973) that had information on contacts with suspected mpox cases had exposures to more than one case. We define the likelihood of a patient with multiple contacts, $\tilde{L}_i(\theta)$, as:

$$\tilde{L}_i(\theta) = \sum_k^{K_i} p_k L_{i,k}(\theta),$$

where $L_{i,k}(\theta)$ is the likelihood for contact $k$ accounting for censoring and truncation in eq. (2), $K_i$ is the total number of reported contacts by patient $i$, and $p_k$ is the probability of each contact being the infector, which we here assume to be uniform ($p_k = 1/K$).

##### S2.2.3 Priors

We inform the model with prior estimates of the incubation period for the 2022 Mpox outbreak in Ponce et al. [2] that found a median incubation period of 9 days, and we set a prior on $\xi$ that puts around 75% of probability mass to values lower than 0.4:

$$\log(\mu) \sim N(\log(9), 0.3),$$
$$\sigma \sim N^+(0, 1),$$
$$\text{logit}(\xi) \sim N(-1, 1).$$

#### S2.3 Model v1: Community infections

As the Mpox epidemic was ongoing in Uvira during the study period there is a chance that infections occurred in the community rather than through reported contacts.

To account for this possibility we need to have priors on when infections may have occurred, which we can account for by jointly modeling the infections curve resulting in the observed epicurve.

##### S2.3.1 Modeling Mpox infections in the community

We here follow methods typically used to infer the basic reproduction number as implemented in the `EpiNow2` R package.

In `EpiNow2` the target of inference is the basic reproduction number, and the package takes as inputs the generation interval and reporting delay distributions including the incubation period and care seeking delays. Here our target of inference is the incubation period itself, and changes in infections are only auxiliary variables used to compute relative probabilities of community infections prior to each reported date of symptom onset.

Following the `EpiNow2` approach, if $\mathcal{I}(t)$ is the true number of infections at time $t$, and knowing the composite delay distribution from infection to reporting $f_D$, we have that the number of modeled reported cases at time $t'$, $y(t')$ is the convolution of the delay distribution and true infections:

$$C(t') = \int_0^{T_D} f_D(\tau)\mathcal{I}(t' - \tau)d\tau,$$

where $T_D$ is the upper bound of the delay distribution.

In our case the delay distribution is the composite of the incubation period and care seeking delay, which can also be written as a convolution:

$$f_D(\tau) = \int_0^{T_R} f_R(\tau')f_I(\tau - \tau')d\tau',$$

where $T_R$ is the upper bound of the care seeking delay distribution, and $f_R$ is the care seeking delay distribution.

The care seeking delay distribution can be informed by the data as we have the reported delays between symptom onset and hospital admission for each patient, $\delta_{R,i}$:

$$\delta_{R,i} \sim f_R(\mu_R, \sigma_R),$$

where $\mu_R$ and $\sigma_R$ are the location and scale of $f_D$. As a first pass we take $f_D$ to be a log-normal distribution.

We can then link the observed and modeled number of cases:

$$y(t) \sim NegBinom(C(t), \eta),$$

where $\eta$ is the overdispersion parameter of the negative binomial distribution.

Assuming that each patient has a probability $\lambda_i$ that each reported contact is the infection contact, the likelihood of a patient's symptom onset date can account for community infections:

$$L(s_i) = \lambda_i' L_{contact}(s_i) + (1 - \lambda_i')L_{comm}(s_i),$$

where $L_{contact}(s_i)$ is the likelihood for contacts as in model v0, $L_{comm}(s_i)$ is the likelihood of the incubation period if infection occurred in the community, and $\lambda_i'$ is the probability that the infection occurred in one of the reported contacts:

$$\lambda_i' = 1 - \prod^{K_i}(1 - \lambda_i).$$

To compute the community likelihood we marginalize out the unknown infection date as:

$$\begin{aligned} L_{comm}(s_i) &= \sum_{t=t_{S_i}-T}^{t_{S_i}} p(t) l_i(t), \\ &= \sum_{t=t_{S_i}-T}^{t_{S_i}} p(t) f_I(t_{S_i} - t), \end{aligned}$$

where $T$ is the assumed upper bound of the incubation period, and $p(t)$ is the probability that community in the infection occurred at time t, and is given by the proportion of the total latent infections in the past $T$ days that occurred on day $t$ as:

$$p(t) = \frac{\mathcal{I}(t)}{\sum_{t'=t_{S_i}-T}^{t_{S_i}} \mathcal{I}(t')}.$$

### S3 Analysis of contact pairs

As a sub-analysis we quantify the incubation period for the subset of cases for which contacts were also enrolled in our study, and for which a date of symptom onset is known. In our dataset we were able to identify 50 cases for which contact pair information was available, along with date of any symptom or rash onset, of which only 41 also reported on date of fever onset (new Supplementary Table S8). Among the 50 cases with contact pair information and date of rash onset, 19 had PCR-positive results and 7 had Ct values below 34, a threshold that we now use to define the high-confidence positive cases, and only one individual reported sexual exposure.

Supplementary Table S8 shows the characteristics of cases retained in the sub-analysis, and Supplementary Figure S6 shows overall and stratified estimates. We did not perform a stratified analysis on exposure type due to limitations in sample size (Supplementary Table S8). Quantile estimates for all distributions and symptom types are provided in Supplementary Table S9.

### S4 Supplementary Figures

#### S4.1 Figure S2: Epidemic curve and timing of patient exposures

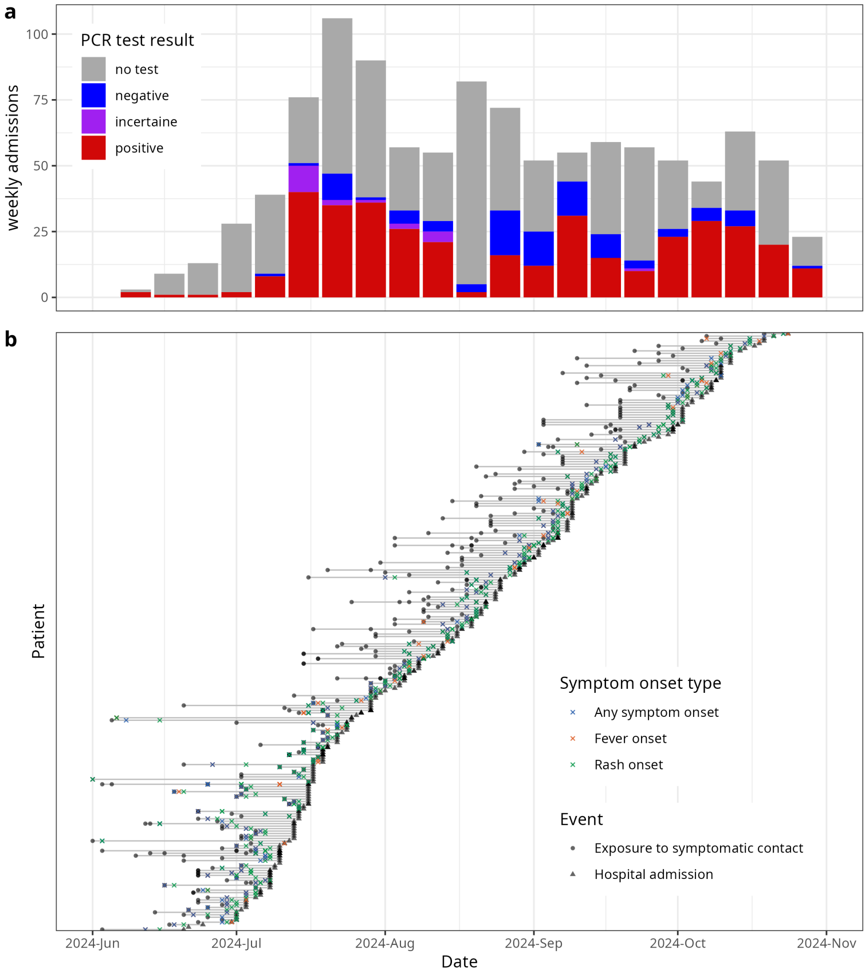

Figure S2: Epidemic curve and timing of patient exposures. a) Mpox epidemic curve in Uvira, DRC, by PCR test result. b) Timing of exposures and symptom onsets of patients with information on symptomatic contacts.

#### S4.2 Figure S3: Distribution of reported times from exposures to symptom onset

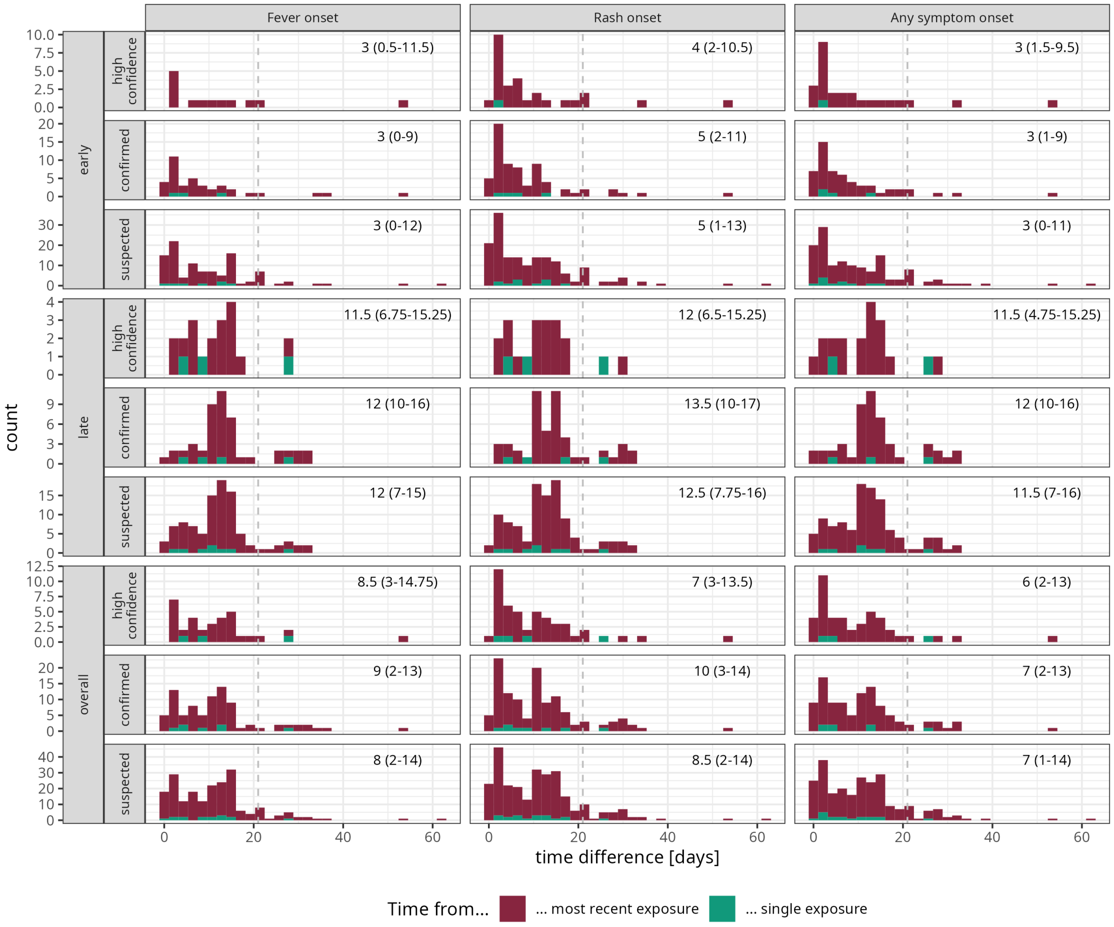

Figure S3: Distribution of reported times from exposures to symptom onset. Time are either to the date of most recent exposure in the case of multiple exposure per contact, or time to single exposure when only one exposure was reported. Vertical lines indicate the three week recall period in the questionnaire (see Questionnaire data). Distributions are shown by epidemic period (early/late: before or after Sep 14 2024) and overall, and by case definition. Numbers indicate the median and inter-quartile range of the distribution.

#### S4.3 Figure S4: Median incubation periods by symptom type

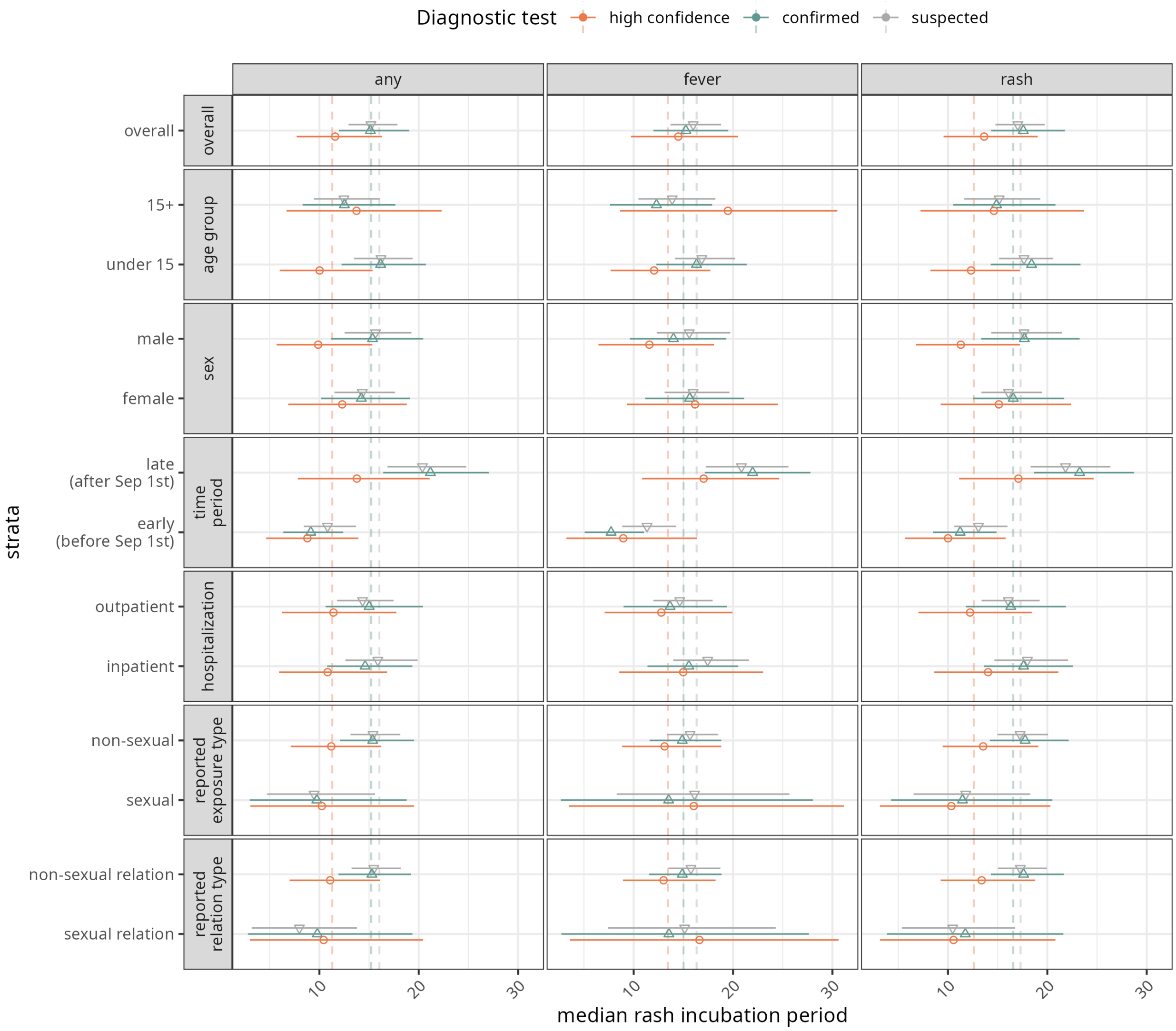

Figure S4: Comparison of median incubation periods by symptom type. Legend as in Figure 1b. Estimates are shown for the main model version (no community infections) and the log-normal distribution. Estimates are given by symptom type (columns), points indicate the mean of 4000 HMC posterior draws and errobars the 95% CrIs.

#### S4.4 Figure S5: Incubation period estimates for varying Ct threholds

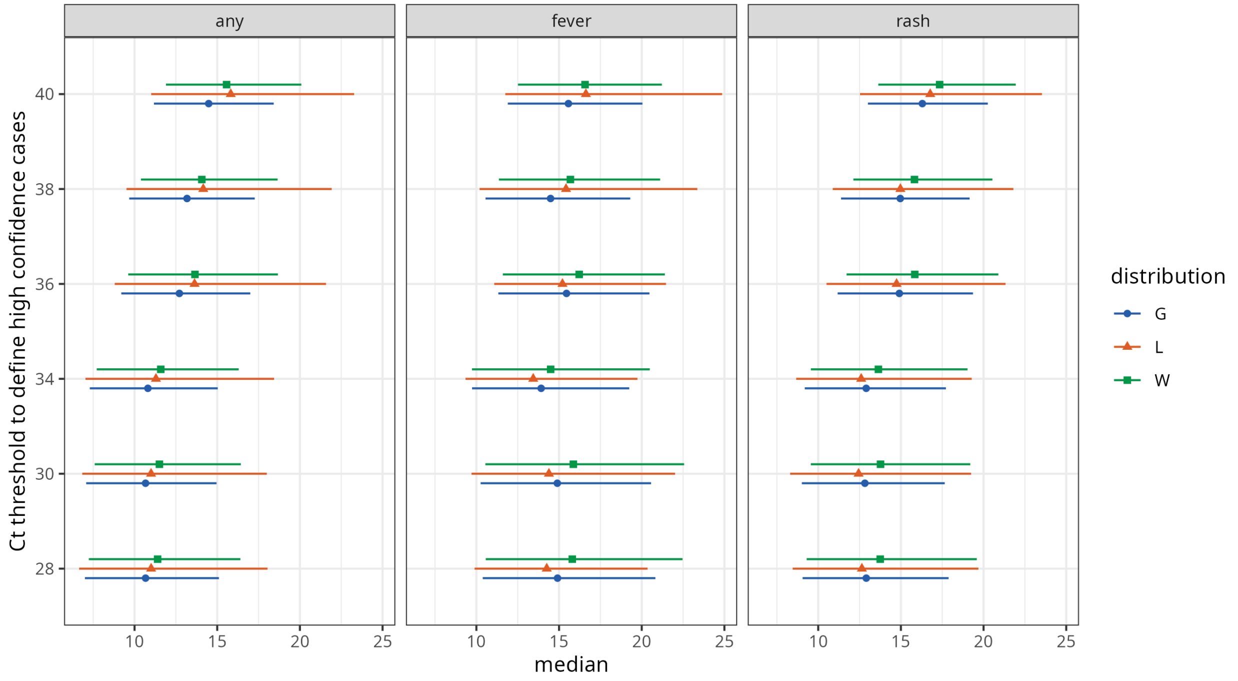

Figure S5: Comparison of incubation period quantiles for high confidence using alternative Ct thresholds. Estimates are given by symptom type (columns), points indicate the mean of 4000 HMC posterior draws and errobars the 95% CrIs.

#### S4.5 Figure S6: Incubation period estimates for subset of contact pairs

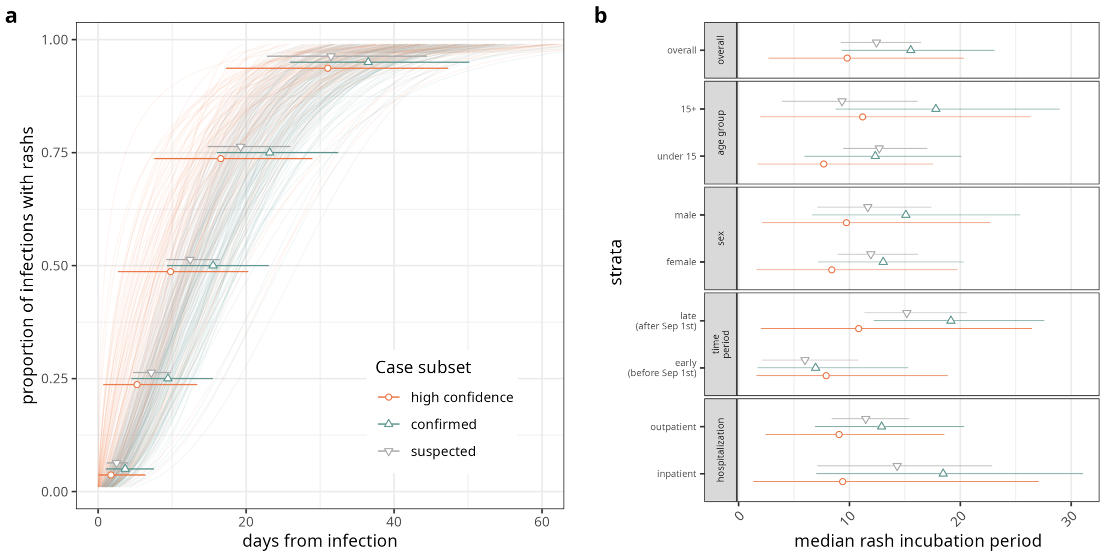

Figure S6: Incubation period quantiles and median estimates by grouping for subset of cases for which contact pairs were also enrolled in the study. Sample sizes given in Supplementary Table S8, and incubation period quantile estimates are given in Supplementary Table S9.

#### S4.6 Figure S7: Comparison with clade 2b and historical incubation period estimates

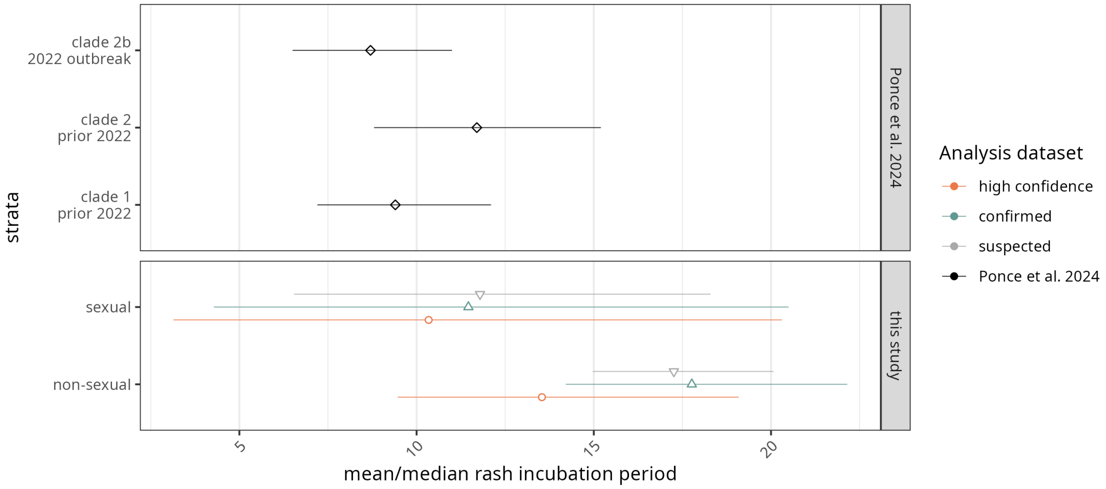

Figure S7: Comparison of median incubation periods in this study and systematic review estimates in [2]. Note that estimates in [2] correspond to the mean incubation period.

### S5 Supplementary Tables

#### S5.1 Table S1: Model comparison results.

Table S1: Model comparison.

| Case definition | Symptom type | Model | Dist | ELPD diff | SE | Rank | Significant | Weight |
|---|---|---|---|---|---|---|---|---|
| **high confidence** | any | v0 | W | 0.0 | 0.0 | 1 | FALSE | 1.000 |
| **high confidence** | any | v0 | G | -0.7 | 0.5 | 2 | FALSE | 0.000 |
| **high confidence** | any | v1 | W | -2.1 | 0.2 | 3 | TRUE | 0.000 |
| **high confidence** | any | v1 | G | -2.6 | 0.3 | 4 | TRUE | 0.000 |
| **high confidence** | any | v0 | L | -2.9 | 2.0 | 5 | FALSE | 0.000 |
| **high confidence** | any | v1 | L | -4.7 | 1.4 | 6 | TRUE | 0.000 |
| **high confidence** | fever | v0 | G | 0.0 | 0.0 | 1 | FALSE | 0.550 |
| **high confidence** | fever | v0 | W | -0.1 | 0.3 | 2 | FALSE | 0.450 |
| **high confidence** | fever | v0 | L | -0.6 | 0.6 | 3 | FALSE | 0.000 |
| **high confidence** | fever | v1 | G | -1.7 | 0.2 | 4 | TRUE | 0.000 |
| **high confidence** | fever | v1 | W | -1.9 | 0.4 | 5 | TRUE | 0.000 |
| **high confidence** | fever | v1 | L | -2.3 | 0.4 | 6 | TRUE | 0.000 |
| **high confidence** | rash | v0 | W | 0.0 | 0.0 | 1 | FALSE | 1.000 |
| **high confidence** | rash | v0 | G | -0.2 | 0.5 | 2 | FALSE | 0.000 |
| **high confidence** | rash | v0 | L | -1.3 | 1.2 | 3 | FALSE | 0.000 |
| **high confidence** | rash | v1 | W | -2.2 | 0.1 | 4 | TRUE | 0.000 |
| **high confidence** | rash | v1 | G | -2.3 | 0.4 | 5 | TRUE | 0.000 |
| **high confidence** | rash | v1 | L | -3.4 | 1.1 | 6 | TRUE | 0.000 |
| **confirmed** | any | v0 | W | 0.0 | 0.0 | 1 | FALSE | 1.000 |
| **confirmed** | any | v0 | G | -1.3 | 0.9 | 2 | FALSE | 0.000 |
| **confirmed** | any | v1 | W | -4.0 | 0.2 | 3 | TRUE | 0.000 |
| **confirmed** | any | v1 | G | -5.3 | 0.6 | 4 | TRUE | 0.000 |
| **confirmed** | any | v0 | L | -5.5 | 3.0 | 5 | FALSE | 0.000 |
| **confirmed** | any | v1 | L | -8.4 | 1.9 | 6 | TRUE | 0.000 |
| **confirmed** | fever | v0 | W | 0.0 | 0.0 | 1 | FALSE | 1.000 |
| **confirmed** | fever | v0 | G | -1.2 | 1.2 | 2 | FALSE | 0.000 |
| **confirmed** | fever | v1 | W | -3.6 | 0.3 | 3 | TRUE | 0.000 |
| **confirmed** | fever | v1 | G | -4.2 | 0.6 | 4 | TRUE | 0.000 |
| **confirmed** | fever | v0 | L | -4.9 | 3.9 | 5 | FALSE | 0.000 |
| **confirmed** | fever | v1 | L | -5.5 | 1.7 | 6 | TRUE | 0.000 |
| **confirmed** | rash | v0 | W | 0.0 | 0.0 | 1 | FALSE | 0.715 |
| **confirmed** | rash | v0 | G | -1.1 | 0.9 | 2 | FALSE | 0.284 |
| **confirmed** | rash | v0 | L | -3.6 | 2.0 | 3 | FALSE | 0.001 |
| **confirmed** | rash | v1 | W | -4.1 | 0.1 | 4 | TRUE | 0.000 |
| **confirmed** | rash | v1 | G | -5.1 | 0.8 | 5 | TRUE | 0.000 |
| **confirmed** | rash | v1 | L | -7.6 | 1.8 | 6 | TRUE | 0.000 |
| **suspected** | any | v0 | W | 0.0 | 0.0 | 1 | FALSE | 1.000 |
| **suspected** | any | v0 | G | -2.0 | 1.0 | 2 | TRUE | 0.000 |
| **suspected** | any | v1 | W | -6.8 | 0.2 | 3 | TRUE | 0.000 |
| **suspected** | any | v1 | G | -8.8 | 0.9 | 4 | TRUE | 0.000 |
| **suspected** | any | v0 | L | -9.6 | 3.3 | 5 | TRUE | 0.000 |
| **suspected** | any | v1 | L | -16.0 | 2.9 | 6 | TRUE | 0.000 |
| **suspected** | fever | v0 | W | 0.0 | 0.0 | 1 | FALSE | 0.623 |
| **suspected** | fever | v0 | G | -1.5 | 1.4 | 2 | FALSE | 0.376 |
| **suspected** | fever | v1 | W | -6.0 | 0.3 | 3 | TRUE | 0.000 |
| **suspected** | fever | v0 | L | -7.1 | 4.5 | 4 | FALSE | 0.000 |
| **suspected** | fever | v1 | G | -7.3 | 1.2 | 5 | TRUE | 0.000 |
| **suspected** | fever | v1 | L | -10.3 | 3.0 | 6 | TRUE | 0.000 |
| **suspected** | rash | v0 | W | 0.0 | 0.0 | 1 | FALSE | 0.999 |
| **suspected** | rash | v0 | G | -2.5 | 1.3 | 2 | FALSE | 0.001 |
| **suspected** | rash | v1 | W | -6.8 | 0.2 | 3 | TRUE | 0.000 |
| **suspected** | rash | v1 | G | -9.1 | 1.1 | 4 | TRUE | 0.000 |
| **suspected** | rash | v0 | L | -10.1 | 4.1 | 5 | TRUE | 0.000 |
| **suspected** | rash | v1 | L | -14.9 | 2.7 | 6 | TRUE | 0.000 |

#### S5.2 Table S2: Incubation quantile estimates for all models

Table S2: Quantile estimates for all models. G: Gamma distribution, L: log-nomral, W: Weibull.

| | | | Quantile | | |
|---|---|---|---|---|---|
| Case definition | Symptom type | Dist | 5% | 50% | 95% |
| **high confidence** | any | G | 1.9 (0.6-3.7) | 10.8 (7.3-15.0) | 34.3 (24.3-46.8) |
| **high confidence** | any | L | 2.3 (1.1-3.8) | 11.3 (7.0-18.4) | 60.5 (27.3-141.7) |
| **high confidence** | any | W | 1.9 (0.7-3.8) | 11.6 (7.7-16.3) | 33.2 (22.9-46.5) |
| **high confidence** | fever | G | 4.4 (1.7-7.6) | 13.9 (9.7-19.2) | 32.8 (23.0-46.1) |
| **high confidence** | fever | L | 5.1 (2.7-8.0) | 13.4 (9.3-19.7) | 37.4 (21.4-76.3) |
| **high confidence** | fever | W | 3.8 (1.3-7.2) | 14.5 (9.7-20.5) | 31.9 (22.7-45.0) |
| **high confidence** | rash | G | 3.3 (1.5-5.7) | 12.9 (9.2-17.7) | 33.7 (23.4-47.2) |
| **high confidence** | rash | L | 3.8 (2.2-5.7) | 12.6 (8.7-19.3) | 44.4 (23.9-92.0) |
| **high confidence** | rash | W | 3.1 (1.3-5.5) | 13.6 (9.6-19.0) | 32.3 (22.4-45.8) |
| **confirmed** | any | G | 3.3 (1.9-4.8) | 14.3 (11.4-17.8) | 39.3 (30.6-49.8) |
| **confirmed** | any | L | 3.6 (2.4-4.9) | 15.2 (11.2-21.6) | 67.3 (38.5-122.2) |
| **confirmed** | any | W | 3.2 (1.8-4.9) | 15.1 (12.0-19.0) | 36.8 (28.7-47.5) |
| **confirmed** | fever | G | 3.6 (2.0-5.3) | 14.4 (11.4-18.1) | 38.0 (29.2-48.8) |
| **confirmed** | fever | L | 3.9 (2.6-5.3) | 15.0 (10.9-21.7) | 60.2 (35.0-111.8) |
| **confirmed** | fever | W | 3.4 (2.0-5.2) | 15.3 (12.0-19.5) | 36.1 (27.9-47.1) |
| **confirmed** | rash | G | 5.0 (3.3-6.8) | 16.5 (13.5-20.2) | 39.4 (31.1-50.4) |
| **confirmed** | rash | L | 5.2 (3.8-6.9) | 16.6 (12.8-22.0) | 53.4 (34.3-86.6) |
| **confirmed** | rash | W | 4.8 (3.0-6.8) | 17.6 (14.3-21.8) | 37.1 (29.6-47.0) |
| **suspected** | any | G | 3.0 (2.1-4.0) | 14.7 (12.5-17.4) | 42.5 (35.0-51.9) |
| **suspected** | any | L | 3.4 (2.6-4.2) | 16.0 (12.7-20.8) | 77.7 (50.5-124.4) |
| **suspected** | any | W | 2.9 (2.0-3.8) | 15.2 (12.9-17.9) | 38.9 (32.5-47.7) |
| **suspected** | fever | G | 4.2 (3.0-5.4) | 15.5 (13.1-18.4) | 39.1 (31.7-48.9) |
| **suspected** | fever | L | 4.5 (3.5-5.6) | 16.3 (13.3-20.7) | 60.3 (40.2-94.2) |
| **suspected** | fever | W | 3.9 (2.8-5.1) | 16.0 (13.7-18.8) | 35.7 (29.6-44.0) |
| **suspected** | rash | G | 4.3 (3.3-5.3) | 16.4 (14.3-18.9) | 42.0 (35.1-50.3) |
| **suspected** | rash | L | 4.6 (3.7-5.6) | 17.3 (14.1-21.5) | 65.7 (45.9-96.6) |
| **suspected** | rash | W | 4.1 (3.1-5.3) | 17.0 (14.8-19.7) | 38.1 (32.1-45.8) |

#### S5.3 Tables S3-S5: Stratified estimates of median incubation period

Table S3: Median incubation period estimates by stratification and symptom type. Estimates shwon for model version v0 and PCR-confirmed cases with Ct values below 34 with contact information.

| | | | | Symptom type | | |
|---|---|---|---|---|---|---|
| Case definition | Grouping | Strata | Dist | Any symptom | Fever | Rash |
| **high confidence** | age group | under 15 | G | 11.8 (8.2-16.4) | 11.6 (7.6-16.5) | 9.5 (5.8-14.0) |
| **high confidence** | age group | under 15 | L | 11.3 (7.8-17.0) | 11.1 (7.7-16.5) | 9.8 (5.8-16.8) |
| **high confidence** | age group | under 15 | W | 12.3 (8.2-17.3) | 12.1 (7.7-17.7) | 10.0 (6.0-15.4) |
| **high confidence** | age group | 15+ | G | 13.0 (6.4-20.4) | 18.5 (8.6-28.6) | 12.3 (6.0-19.4) |
| **high confidence** | age group | 15+ | L | 13.4 (7.3-23.3) | 18.2 (10.1-28.5) | 12.9 (7.0-22.2) |
| **high confidence** | age group | 15+ | W | 14.6 (7.2-23.7) | 19.5 (8.6-30.5) | 13.7 (6.7-22.3) |
| **high confidence** | sex | female | G | 14.0 (9.0-20.1) | 15.0 (9.3-22.0) | 10.9 (5.9-16.4) |
| **high confidence** | sex | female | L | 13.9 (8.8-21.9) | 14.9 (9.2-23.5) | 12.2 (6.6-21.7) |
| **high confidence** | sex | female | W | 15.1 (9.3-22.4) | 16.2 (9.3-24.5) | 12.3 (6.9-18.8) |
| **high confidence** | sex | male | G | 10.8 (6.7-16.3) | 11.6 (6.5-17.7) | 9.6 (5.5-14.6) |
| **high confidence** | sex | male | L | 10.3 (6.5-17.3) | 11.5 (7.1-16.5) | 9.2 (5.7-15.5) |
| **high confidence** | sex | male | W | 11.3 (6.8-17.2) | 11.6 (6.5-18.1) | 9.9 (5.7-15.3) |
| **high confidence** | time period | early (before Sep 1st) | G | 9.7 (5.8-14.4) | 9.1 (3.8-15.7) | 8.7 (4.9-13.1) |
| **high confidence** | time period | early (before Sep 1st) | L | 9.0 (5.5-14.3) | 9.3 (5.2-14.3) | 8.4 (5.0-13.9) |
| **high confidence** | time period | early (before Sep 1st) | W | 10.0 (5.7-15.8) | 9.0 (3.2-16.4) | 8.8 (4.6-13.9) |
| **high confidence** | time period | late (after Sep 1st) | G | 16.0 (10.6-22.5) | 15.7 (10.3-22.3) | 12.0 (7.1-18.0) |
| **high confidence** | time period | late (after Sep 1st) | L | 15.8 (10.4-24.6) | 15.6 (10.2-24.6) | 13.6 (7.4-24.7) |
| **high confidence** | time period | late (after Sep 1st) | W | 17.1 (11.1-24.7) | 17.0 (10.8-24.7) | 13.8 (7.8-21.1) |
| **high confidence** | hospitalization | inpatient | G | 13.0 (8.2-18.4) | 14.0 (8.4-20.4) | 10.0 (5.7-15.1) |
| **high confidence** | hospitalization | inpatient | L | 12.8 (7.8-21.2) | 13.6 (8.5-21.8) | 10.8 (5.9-19.3) |
| **high confidence** | hospitalization | inpatient | W | 14.0 (8.6-21.1) | 15.0 (8.5-23.0) | 10.8 (5.9-16.8) |
| **high confidence** | hospitalization | outpatient | G | 11.9 (6.9-17.7) | 12.7 (7.0-19.3) | 11.2 (6.3-17.0) |
| **high confidence** | hospitalization | outpatient | L | 11.8 (7.4-18.4) | 13.1 (8.0-19.4) | 11.2 (6.9-17.6) |
| **high confidence** | hospitalization | outpatient | W | 12.2 (7.0-18.4) | 12.8 (7.1-19.9) | 11.4 (6.2-17.7) |
| **high confidence** | reported exposure type | sexual | G | 9.5 (3.1-17.0) | 15.7 (2.4-30.7) | 9.3 (2.9-16.8) |
| **high confidence** | reported exposure type | sexual | L | 10.7 (5.1-20.4) | 18.5 (7.4-31.0) | 10.7 (5.1-20.2) |
| **high confidence** | reported exposure type | sexual | W | 10.3 (3.1-20.3) | 16.0 (3.5-31.2) | 10.2 (3.1-19.5) |
| **high confidence** | reported exposure type | non-sexual | G | 13.0 (9.1-17.8) | 12.7 (8.6-17.8) | 10.5 (6.9-15.1) |
| **high confidence** | reported exposure type | non-sexual | L | 12.4 (8.8-18.3) | 12.0 (8.2-17.2) | 10.9 (6.8-17.9) |
| **high confidence** | reported exposure type | non-sexual | W | 13.5 (9.5-19.1) | 13.1 (8.8-18.8) | 11.2 (7.1-16.2) |
| **high confidence** | reported relation type | sexual relation | G | 9.2 (2.4-17.2) | 15.4 (2.3-30.8) | 9.3 (2.2-17.4) |
| **high confidence** | reported relation type | sexual relation | L | 11.0 (5.0-21.4) | 18.3 (7.5-31.3) | 11.1 (5.3-21.5) |
| **high confidence** | reported relation type | sexual relation | W | 10.6 (3.2-20.8) | 16.6 (3.6-30.6) | 10.4 (3.0-20.4) |
| **high confidence** | reported relation type | non-sexual relation | G | 12.8 (9.2-17.5) | 12.7 (8.6-17.8) | 10.5 (6.8-15.0) |
| **high confidence** | reported relation type | non-sexual relation | L | 12.2 (8.7-17.8) | 12.1 (8.4-17.7) | 10.8 (6.7-17.8) |
| **high confidence** | reported relation type | non-sexual relation | W | 13.4 (9.3-18.8) | 13.0 (8.9-18.2) | 11.1 (7.0-16.1) |

Table S4: Median incubation period estimates by stratification and symptom type. Estimates shwon for model version v0 and PCR-confirmed cases with contact information.

| Case definition | Grouping | Strata | Dist | Symptom type | | |
|---|---|---|---|---|---|---|
| | | | | Any symptom | Fever | Rash |
| **confirmed** | age group | under 15 | G | 17.3 (13.8-21.6) | 15.2 (11.5-19.6) | 15.0 (11.5-19.4) |
| **confirmed** | age group | under 15 | L | 17.4 (13.2-23.6) | 16.4 (11.3-24.9) | 16.4 (11.4-24.4) |
| **confirmed** | age group | under 15 | W | 18.4 (14.3-23.3) | 16.3 (12.3-21.4) | 16.2 (12.2-20.7) |
| **confirmed** | age group | 15+ | G | 13.9 (9.9-19.1) | 11.7 (7.3-16.6) | 11.8 (8.0-16.5) |
| **confirmed** | age group | 15+ | L | 14.1 (9.4-21.8) | 11.7 (7.5-18.0) | 12.0 (7.7-19.1) |
| **confirmed** | age group | 15+ | W | 14.9 (10.5-20.8) | 12.3 (7.6-17.9) | 12.5 (8.3-17.6) |
| **confirmed** | sex | female | G | 15.6 (12.0-20.1) | 14.8 (11.1-19.4) | 13.3 (9.8-17.4) |
| **confirmed** | sex | female | L | 15.3 (11.6-20.5) | 14.5 (10.8-20.6) | 14.3 (9.5-21.7) |
| **confirmed** | sex | female | W | 16.6 (12.5-21.7) | 15.6 (11.2-21.1) | 14.2 (10.2-19.1) |
| **confirmed** | sex | male | G | 16.3 (12.4-21.3) | 12.9 (9.0-17.5) | 14.3 (10.4-18.8) |
| **confirmed** | sex | male | L | 17.1 (12.1-25.4) | 14.0 (8.9-22.1) | 15.0 (10.4-22.2) |
| **confirmed** | sex | male | W | 17.7 (13.3-23.2) | 14.0 (9.6-19.3) | 15.3 (11.2-20.4) |
| **confirmed** | time period | early (before Sep 1st) | G | 10.9 (8.2-14.5) | 7.6 (4.9-10.9) | 8.9 (6.4-12.0) |
| **confirmed** | time period | early (before Sep 1st) | L | 10.4 (7.6-14.6) | 7.2 (4.9-10.9) | 8.5 (6.0-12.1) |
| **confirmed** | time period | early (before Sep 1st) | W | 11.2 (8.5-14.9) | 7.7 (5.1-11.0) | 9.1 (6.4-12.4) |
| **confirmed** | time period | late (after Sep 1st) | G | 21.6 (17.5-26.6) | 20.2 (15.9-25.5) | 19.3 (14.8-24.6) |
| **confirmed** | time period | late (after Sep 1st) | L | 21.3 (16.8-28.0) | 19.9 (15.5-26.8) | 21.8 (14.9-32.7) |
| **confirmed** | time period | late (after Sep 1st) | W | 23.3 (18.7-28.7) | 22.0 (17.2-27.8) | 21.2 (16.4-27.1) |
| **confirmed** | hospitalization | inpatient | G | 16.5 (12.9-21.2) | 14.6 (11.0-19.1) | 13.7 (10.2-17.9) |
| **confirmed** | hospitalization | inpatient | L | 16.6 (12.3-23.4) | 14.3 (10.6-20.0) | 14.9 (9.9-23.5) |
| **confirmed** | hospitalization | inpatient | W | 17.6 (13.6-22.6) | 15.6 (11.4-20.5) | 14.6 (10.8-19.4) |
| **confirmed** | hospitalization | outpatient | G | 15.3 (11.2-20.1) | 12.5 (8.3-17.3) | 14.0 (10.1-18.6) |
| **confirmed** | hospitalization | outpatient | L | 15.7 (10.8-23.5) | 13.7 (8.6-21.7) | 14.5 (9.9-21.4) |
| **confirmed** | hospitalization | outpatient | W | 16.3 (11.8-21.9) | 13.7 (9.0-19.4) | 15.0 (10.6-20.4) |
| **confirmed** | reported exposure type | sexual | G | 10.3 (4.0-18.2) | 13.0 (1.8-27.3) | 9.0 (2.8-16.4) |
| **confirmed** | reported exposure type | sexual | L | 11.4 (5.7-21.8) | 15.6 (6.4-28.6) | 10.5 (4.9-19.7) |
| **confirmed** | reported exposure type | sexual | W | 11.5 (4.3-20.5) | 13.5 (2.7-28.0) | 9.7 (3.0-18.8) |
| **confirmed** | reported exposure type | non-sexual | G | 16.7 (13.6-20.5) | 14.1 (11.2-17.8) | 14.6 (11.6-18.1) |
| **confirmed** | reported exposure type | non-sexual | L | 16.6 (13.0-22.1) | 14.6 (10.8-20.4) | 15.5 (11.3-21.8) |
| **confirmed** | reported exposure type | non-sexual | W | 17.8 (14.2-22.1) | 14.9 (11.6-18.8) | 15.4 (12.1-19.5) |
| **confirmed** | reported relation type | sexual relation | G | 10.3 (3.4-18.3) | 12.8 (1.9-27.2) | 8.8 (2.3-16.5) |
| **confirmed** | reported relation type | sexual relation | L | 11.8 (5.8-22.1) | 15.8 (6.7-29.1) | 10.8 (5.2-21.0) |
| **confirmed** | reported relation type | sexual relation | W | 11.7 (3.9-21.6) | 13.5 (2.7-27.6) | 9.8 (2.8-19.4) |
| **confirmed** | reported relation type | non-sexual relation | G | 16.6 (13.5-20.3) | 14.1 (11.1-17.8) | 14.5 (11.6-18.1) |
| **confirmed** | reported relation type | non-sexual relation | L | 16.5 (13.0-21.9) | 14.8 (10.7-21.2) | 15.4 (11.3-22.2) |
| **confirmed** | reported relation type | non-sexual relation | W | 17.6 (14.3-21.7) | 14.9 (11.6-18.8) | 15.3 (11.9-19.2) |

Table S5: Median incubation period estimates by stratification and symptom type. Estimates shwon for model version v0 and suspected with contact information.

| | | | | Symptom type | | |
|---|---|---|---|---|---|---|
| Case definition | Grouping | Strata | Dist | Any symptom | Fever | Rash |
| **suspected** | age group | under 15 | G | 16.9 (14.6-19.9) | 16.2 (13.6-19.3) | 15.6 (13.2-18.8) |
| **suspected** | age group | under 15 | L | 16.8 (14.2-20.3) | 16.8 (13.4-21.8) | 16.5 (13.1-21.6) |
| **suspected** | age group | under 15 | W | 17.6 (15.1-20.6) | 16.9 (14.2-20.2) | 16.2 (13.5-19.4) |
| **suspected** | age group | 15+ | G | 14.2 (11.1-18.0) | 13.2 (10.0-17.1) | 11.9 (9.0-15.1) |
| **suspected** | age group | 15+ | L | 15.9 (11.0-24.1) | 14.0 (9.9-20.9) | 13.2 (9.1-20.0) |
| **suspected** | age group | 15+ | W | 15.1 (11.6-19.3) | 13.9 (10.5-18.2) | 12.5 (9.4-16.1) |
| **suspected** | sex | female | G | 15.6 (12.9-18.7) | 15.5 (12.8-18.9) | 13.8 (11.1-17.0) |
| **suspected** | sex | female | L | 16.6 (12.9-22.3) | 15.9 (12.6-20.9) | 15.8 (11.4-22.2) |
| **suspected** | sex | female | W | 16.1 (13.4-19.5) | 16.0 (13.1-19.6) | 14.3 (11.5-17.6) |
| **suspected** | sex | male | G | 16.7 (13.6-20.2) | 14.8 (11.8-18.4) | 14.9 (12.1-18.2) |
| **suspected** | sex | male | L | 17.5 (13.7-23.3) | 15.6 (11.5-21.8) | 15.5 (11.8-21.1) |
| **suspected** | sex | male | W | 17.7 (14.4-21.5) | 15.6 (12.3-19.7) | 15.6 (12.5-19.3) |
| **suspected** | time period | early (before Sep 1st) | G | 12.7 (10.4-15.3) | 11.0 (8.6-13.9) | 10.5 (8.4-13.1) |
| **suspected** | time period | early (before Sep 1st) | L | 12.9 (10.0-17.2) | 11.4 (8.4-16.4) | 10.8 (8.2-14.8) |
| **suspected** | time period | early (before Sep 1st) | W | 13.1 (10.6-16.0) | 11.3 (8.8-14.3) | 10.8 (8.4-13.7) |
| **suspected** | time period | late (after Sep 1st) | G | 20.6 (17.3-24.8) | 19.5 (16.3-23.7) | 19.2 (16.0-23.2) |
| **suspected** | time period | late (after Sep 1st) | L | 21.7 (17.2-28.7) | 19.6 (15.7-25.5) | 21.5 (16.2-29.4) |
| **suspected** | time period | late (after Sep 1st) | W | 21.8 (18.3-26.3) | 20.8 (17.3-25.6) | 20.4 (16.9-24.8) |
| **suspected** | hospitalization | inpatient | G | 16.9 (13.8-20.7) | 16.5 (13.3-20.4) | 14.8 (11.8-18.6) |
| **suspected** | hospitalization | inpatient | L | 18.5 (13.8-25.5) | 17.6 (13.1-24.9) | 17.6 (12.4-26.4) |
| **suspected** | hospitalization | inpatient | W | 18.0 (14.7-22.1) | 17.4 (14.0-21.6) | 15.9 (12.6-19.9) |
| **suspected** | hospitalization | outpatient | G | 15.4 (12.9-18.4) | 14.2 (11.6-17.3) | 13.8 (11.3-16.8) |
| **suspected** | hospitalization | outpatient | L | 15.8 (12.7-20.1) | 14.8 (11.4-19.9) | 14.1 (11.1-18.7) |
| **suspected** | hospitalization | outpatient | W | 16.1 (13.4-19.2) | 14.6 (12.0-18.0) | 14.3 (11.8-17.5) |
| **suspected** | reported exposure type | sexual | G | 10.4 (6.1-15.7) | 13.8 (7.0-21.5) | 8.5 (4.1-13.3) |
| **suspected** | reported exposure type | sexual | L | 12.8 (6.9-23.6) | 15.9 (8.3-29.2) | 11.1 (5.6-20.5) |
| **suspected** | reported exposure type | sexual | W | 11.8 (6.5-18.3) | 16.1 (8.3-25.7) | 9.5 (4.7-15.6) |
| **suspected** | reported exposure type | non-sexual | G | 16.6 (14.4-19.3) | 15.2 (12.9-18.0) | 14.9 (12.8-17.5) |
| **suspected** | reported exposure type | non-sexual | L | 16.6 (14.1-20.1) | 15.6 (12.8-19.6) | 15.5 (12.6-19.7) |
| **suspected** | reported exposure type | non-sexual | W | 17.3 (15.0-20.1) | 15.7 (13.4-18.5) | 15.4 (13.1-18.1) |
| **suspected** | reported relation type | sexual relation | G | 9.4 (4.9-14.8) | 12.7 (5.7-20.8) | 7.1 (2.9-12.2) |
| **suspected** | reported relation type | sexual relation | L | 11.7 (6.1-21.4) | 15.1 (7.4-28.5) | 9.9 (4.9-19.5) |
| **suspected** | reported relation type | sexual relation | W | 10.5 (5.4-16.8) | 15.1 (7.4-24.3) | 8.0 (3.2-13.8) |
| **suspected** | reported relation type | non-sexual relation | G | 16.6 (14.4-19.3) | 15.2 (13.0-17.9) | 15.0 (12.8-17.6) |
| **suspected** | reported relation type | non-sexual relation | L | 16.6 (14.1-20.2) | 15.7 (12.9-19.9) | 15.5 (12.7-19.6) |
| **suspected** | reported relation type | non-sexual relation | W | 17.3 (15.0-20.0) | 15.8 (13.5-18.7) | 15.5 (13.2-18.2) |

#### S5.4 Table S6: Probabilities of exceedence

Table S6: Probability of the incubation period exceeding two, three, and four weeks. G: Gamma distribution, L: log-nomral, W: Weibull.

| | | | Delay | | |
|---|---|---|---|---|---|
| Case definition | Symptom | Dist | 14 days | 21 days | 28 days |
| **high confidence** | any | G | 37.3% (22.6-53.8) | 19.3% (8.3-32.9) | 9.8% (2.7-19.8) |
| **high confidence** | any | L | 39.6% (21.6-59.6) | 25.2% (9.8-45.7) | 17.2% (4.7-36.4) |
| **high confidence** | any | W | 40.1% (23.6-58.5) | 20.2% (7.4-35.9) | 9.5% (1.5-21.3) |
| **high confidence** | fever | G | 49.1% (28.8-71.7) | 23.1% (7.8-43.8) | 10.0% (1.4-24.1) |
| **high confidence** | fever | L | 45.5% (23.8-70.5) | 21.8% (5.5-46.5) | 11.1% (1.1-32.3) |
| **high confidence** | fever | W | 51.6% (30.3-74.9) | 24.8% (8.0-48.5) | 9.9% (0.7-26.8) |
| **high confidence** | rash | G | 44.5% (26.7-63.9) | 21.9% (8.0-39.6) | 10.3% (1.9-23.6) |
| **high confidence** | rash | L | 42.6% (23.5-63.8) | 23.5% (7.8-46.0) | 13.9% (2.7-34.1) |
| **high confidence** | rash | W | 47.7% (29.1-68.0) | 23.2% (7.2-43.6) | 9.9% (0.8-25.2) |
| **confirmed** | any | G | 50.9% (39.0-62.7) | 28.7% (17.7-40.4) | 15.3% (7.1-24.9) |
| **confirmed** | any | L | 52.7% (38.7-67.0) | 34.9% (20.8-51.0) | 24.0% (11.5-39.8) |
| **confirmed** | any | W | 54.0% (41.5-67.1) | 30.1% (18.0-43.8) | 14.7% (5.7-26.3) |
| **confirmed** | fever | G | 51.3% (38.6-64.4) | 28.1% (16.5-41.5) | 14.4% (6.1-25.1) |
| **confirmed** | fever | L | 52.2% (37.4-67.6) | 33.2% (18.7-51.3) | 22.0% (9.5-39.5) |
| **confirmed** | fever | W | 54.5% (41.3-68.7) | 29.9% (17.3-45.4) | 14.2% (4.9-27.4) |
| **confirmed** | rash | G | 59.8% (47.6-71.7) | 33.8% (21.4-47.4) | 17.3% (8.0-29.1) |
| **confirmed** | rash | L | 58.6% (44.4-72.3) | 36.0% (21.1-52.4) | 22.3% (9.9-38.3) |
| **confirmed** | rash | W | 63.8% (51.5-76.2) | 37.0% (23.5-52.6) | 17.7% (6.9-32.1) |
| **suspected** | any | G | 52.4% (43.9-60.6) | 31.2% (22.9-40.0) | 17.7% (11.1-25.6) |
| **suspected** | any | L | 55.1% (45.3-64.8) | 38.3% (27.7-49.6) | 27.4% (17.6-39.0) |
| **suspected** | any | W | 54.1% (45.8-62.5) | 31.5% (23.1-41.1) | 16.5% (9.6-25.2) |
| **suspected** | fever | G | 55.8% (45.8-65.3) | 31.3% (21.4-42.2) | 16.1% (8.6-25.7) |
| **suspected** | fever | L | 57.2% (47.0-67.5) | 36.8% (25.4-49.4) | 24.2% (13.7-36.9) |
| **suspected** | fever | W | 57.8% (48.7-67.2) | 31.7% (21.9-42.8) | 14.6% (6.9-24.7) |
| **suspected** | rash | G | 58.9% (51.1-66.7) | 35.0% (26.5-43.9) | 19.2% (12.1-27.3) |
| **suspected** | rash | L | 59.9% (50.5-69.1) | 40.0% (29.6-51.1) | 27.1% (17.3-38.7) |
| **suspected** | rash | W | 61.3% (53.2-69.4) | 36.0% (26.9-46.0) | 18.0% (10.3-27.4) |

#### S5.5 Table S7: Parameter estimates

Table S7: Parameter estimates for each distribution and symptom type for model version v0. Posterior samples are provided as csv files at https://github.com/HopkinsIDD/mpox_cladeIb_incubation_period. Parametrizations follow the Stan distribution definitions at https://mc-stan.org/docs/functions-reference/positive_continuous_distributions.html: Log-normal = {mu, sigma}, Gamma = {alpha, beta}, Weibull = {alpha, sigma}.

| | | Log-normal | | Gamma | | Weibull | |
|---|---|---|---|---|---|---|---|
| Case definition | Symptom | param 1 | param 2 | param 1 | param 2 | param 1 | param 2 |
| **high confidence** | any | 2.4 (2.0-2.9) | 1.0 (0.7-1.4) | 1.7 (1.0-2.7) | 0.1 (0.1-0.2) | 1.4 (1.0-1.9) | 15.0 (10.5-20.7) |
| **high confidence** | fever | 2.6 (2.2-3.0) | 0.6 (0.4-0.9) | 3.3 (1.6-6.4) | 0.2 (0.1-0.4) | 1.9 (1.2-2.9) | 17.6 (12.5-24.4) |
| **high confidence** | rash | 2.5 (2.2-3.0) | 0.7 (0.5-1.0) | 2.5 (1.4-4.3) | 0.2 (0.1-0.3) | 1.8 (1.2-2.5) | 16.9 (12.3-23.3) |
| **confirmed** | any | 2.7 (2.4-3.1) | 0.9 (0.7-1.1) | 2.2 (1.5-3.0) | 0.1 (0.1-0.2) | 1.7 (1.3-2.1) | 18.9 (15.2-23.5) |
| **confirmed** | fever | 2.7 (2.4-3.1) | 0.8 (0.6-1.1) | 2.4 (1.6-3.5) | 0.1 (0.1-0.2) | 1.7 (1.4-2.2) | 18.9 (15.1-24.0) |
| **confirmed** | rash | 2.8 (2.5-3.1) | 0.7 (0.5-0.9) | 3.0 (2.0-4.2) | 0.2 (0.1-0.2) | 2.0 (1.6-2.5) | 21.2 (17.4-26.2) |
| **suspected** | any | 2.8 (2.5-3.0) | 0.9 (0.8-1.1) | 1.9 (1.5-2.4) | 0.1 (0.1-0.2) | 1.6 (1.3-1.8) | 19.2 (16.5-22.7) |
| **suspected** | fever | 2.8 (2.6-3.0) | 0.8 (0.6-0.9) | 2.6 (1.9-3.4) | 0.1 (0.1-0.2) | 1.8 (1.5-2.2) | 19.5 (16.8-23.1) |
| **suspected** | rash | 2.8 (2.6-3.1) | 0.8 (0.7-0.9) | 2.5 (2.0-3.1) | 0.1 (0.1-0.2) | 1.8 (1.6-2.1) | 20.8 (18.1-24.2) |

#### S5.6 Table S8: Sample characteristics of contact pairs subanalysis

Table S8: Characteristics of cases for which contact pairs were also enrolled in the study.

| **Characteristic** | **any**<br>N = 50 | **fever**<br>N = 41 | **rash**<br>N = 50 |
|---|---|---|---|
| Age | | | |
| u15 | 36 (72%) | 28 (68%) | 36 (72%) |
| 15+ | 14 (28%) | 13 (32%) | 14 (28%) |
| Sex | | | |
| female | 27 (54%) | 21 (51%) | 27 (54%) |
| male | 23 (46%) | 20 (49%) | 23 (46%) |
| PCR result | | | |
| positive | 19 (76%) | 18 (82%) | 19 (76%) |
| negative | 6 (24%) | 4 (18%) | 6 (24%) |
| incertaine | 0 (0%) | 0 (0%) | 0 (0%) |
| Unknown | 25 | 19 | 25 |
| Ct category(threshold = 34) | | | |
| low_ct | 7 (41%) | 5 (33%) | 7 (41%) |
| high_ct | 10 (59%) | 10 (67%) | 10 (59%) |
| Unknown | 33 | 26 | 33 |

[1] n (%)

#### S5.7 Table S9: Incubation quantile estimates for contact pair subanalysis

Table S9: Quantile estimates for subanalysis subsetting on contact pairs that were both enrolled in this study. G: Gamma distribution, L: log-nomral, W: Weibull.

| | | | Quantile | | |
|---|---|---|---|---|---|
| Case definition | Symptom type | Dist | 5% | 50% | 95% |
| **high confidence** | any | G | 1.8 (0.0-6.2) | 9.0 (2.5-17.0) | 30.4 (17.1-45.7) |
| **high confidence** | any | L | 3.3 (1.0-7.2) | 9.9 (4.8-19.0) | 34.7 (12.5-96.3) |
| **high confidence** | any | W | 1.7 (0.0-6.4) | 9.6 (2.4-19.5) | 30.5 (17.0-46.6) |
| **high confidence** | fever | G | 4.1 (0.0-12.2) | 12.5 (3.7-23.4) | 31.8 (18.6-48.4) |
| **high confidence** | fever | L | 6.4 (1.5-12.7) | 13.5 (6.1-23.4) | 33.3 (14.8-81.4) |
| **high confidence** | fever | W | 3.4 (0.1-11.3) | 12.8 (3.5-25.6) | 31.5 (17.9-47.7) |
| **high confidence** | rash | G | 1.9 (0.0-6.7) | 9.2 (2.3-18.1) | 30.8 (17.4-46.0) |
| **high confidence** | rash | L | 3.3 (0.9-7.1) | 10.2 (4.9-19.7) | 36.2 (12.9-98.5) |
| **high confidence** | rash | W | 1.7 (0.0-6.4) | 9.8 (2.7-20.3) | 31.0 (17.3-47.3) |
| **confirmed** | any | G | 3.6 (0.9-7.0) | 13.8 (8.5-19.9) | 36.4 (25.1-49.9) |
| **confirmed** | any | L | 4.3 (2.0-7.2) | 14.4 (8.9-23.9) | 52.1 (25.8-117.1) |
| **confirmed** | any | W | 3.3 (0.9-6.8) | 14.9 (9.0-21.8) | 36.1 (25.7-49.3) |
| **confirmed** | fever | G | 2.4 (0.4-5.3) | 12.4 (7.2-18.3) | 37.5 (26.4-50.9) |
| **confirmed** | fever | L | 2.8 (1.2-5.1) | 13.8 (7.9-24.9) | 73.5 (31.0-174.6) |
| **confirmed** | fever | W | 2.7 (0.7-5.8) | 13.8 (8.3-20.5) | 36.3 (25.9-50.0) |
| **confirmed** | rash | G | 3.9 (1.1-7.3) | 14.4 (9.2-20.4) | 37.4 (26.7-51.1) |
| **confirmed** | rash | L | 4.5 (2.0-7.3) | 15.0 (9.4-23.8) | 54.5 (27.3-116.7) |
| **confirmed** | rash | W | 3.7 (1.0-7.5) | 15.5 (9.3-23.1) | 36.5 (25.9-50.2) |
| **suspected** | any | G | 2.6 (1.1-4.2) | 11.5 (8.7-15.0) | 32.4 (23.3-44.7) |
| **suspected** | any | L | 3.3 (2.0-4.7) | 11.6 (8.5-16.9) | 43.1 (24.3-87.3) |
| **suspected** | any | W | 2.1 (1.0-3.6) | 11.7 (8.7-15.6) | 31.5 (23.0-43.4) |
| **suspected** | fever | G | 3.3 (1.5-5.2) | 12.6 (9.4-16.5) | 32.8 (23.3-45.0) |
| **suspected** | fever | L | 3.6 (2.1-5.2) | 13.3 (9.3-20.0) | 51.7 (27.7-106.4) |
| **suspected** | fever | W | 2.9 (1.4-4.6) | 12.8 (9.5-17.0) | 30.4 (22.3-43.1) |
| **suspected** | rash | G | 2.9 (1.3-4.6) | 12.1 (9.2-15.8) | 33.2 (23.8-44.8) |
| **suspected** | rash | L | 3.6 (2.2-5.1) | 12.5 (9.1-18.2) | 45.6 (25.7-92.4) |
| **suspected** | rash | W | 2.5 (1.2-4.0) | 12.4 (9.2-16.5) | 31.5 (22.8-44.4) |

#### S5.8 Table S10: Symptom types by analysis subset

Table S10: Frequency of symptoms by case definition.

| | Case definition | | |
|---|---|---|---|
| Symptom | suspected | confirmed | high-confidence |
| Fever | 197 (81%) | 78 (85%) | 29 (78%) |
| Axillary lymphadenopathy | 86 (35%) | 37 (40%) | 11 (30%) |
| Cervical lymphadenopathy | 163 (67%) | 67 (73%) | 25 (68%) |
| Inguinal lymphadenopathy | 186 (77%) | 72 (78%) | 27 (73%) |
| General lymphadenopathy | 60 (25%) | 29 (32%) | 11 (30%) |
| Hospitalized | 107 (44%) | 52 (57%) | 20 (54%) |
| Skin lesions | 222 (91%) | 83 (90%) | 35 (95%) |
| Palm or sole lesions | 82 (34%) | 37 (40%) | 18 (49%) |
| Anal lesions | 35 (14%) | 14 (15%) | 3 (8%) |
| Genital lesions | 107 (44%) | 45 (49%) | 14 (38%) |
| Oral lesions | 57 (23%) | 27 (29%) | 9 (24%) |
| Cough | 88 (36%) | 38 (41%) | 17 (46%) |
| Sore throat | 106 (44%) | 45 (49%) | 21 (57%) |
| Vomiting/nausea | 46 (19%) | 25 (27%) | 11 (30%) |
| Asthenia | 151 (62%) | 64 (70%) | 23 (62%) |
| Myalgia | 88 (36%) | 37 (40%) | 11 (30%) |
| Conjunctivitis | 7 (3%) | 6 (7%) | 1 (3%) |
| Eye itching | 41 (17%) | 19 (21%) | 6 (16%) |
| Chills/sweats | 151 (62%) | 57 (62%) | 23 (62%) |
| Headache | 115 (47%) | 46 (50%) | 16 (43%) |
| Light sensitivity | 50 (21%) | 23 (25%) | 5 (14%) |
| Confined to bed | 19 (8%) | 13 (14%) | 4 (11%) |
| Diarrhea | 35 (14%) | 18 (20%) | 7 (19%) |
| Rectal pain/bleeding | 8 (3%) | 4 (4%) | 1 (3%) |

##### S5.9 Table S11: Symptom types by exposure type

Table S11: Comparison of symptom frequency between sexually-exposed (N=20) and non-sexually exposed (N=223) cases. 95% confidence intervals (CI) for risk ratio estimates were calculated using the epitools R package [4] with the small sample adjustment. P-values were calculated using the mid-P exact test.

| Symptom | Sexual exposure | Non-sexual exposure | Risk Ratio (9\5% CI) | p-value |
|---|---|---|---|---|
| Fever | 13 (65%) | 184 (83%) | 0.79 (0.57-1.09) | 0.08 |
| Axillary lymphadenopathy | 4 (20%) | 82 (37%) | 0.54 (0.22-1.32) | 0.14 |
| Cervical lymphadenopathy | 7 (35%) | 156 (70%) | 0.5 (0.27-0.91) | 0.001 |
| Inguinal lymphadenopathy | 16 (80%) | 170 (76%) | 1.05 (0.83-1.32) | 0.74 |
| General lymphadenopathy | 2 (10%) | 58 (26%) | 0.38 (0.1-1.44) | 0.11 |
| Hospitalized | 13 (65%) | 94 (42%) | 1.53 (1.07-2.19) | 0.06 |
| Skin lesions | 16 (80%) | 206 (92%) | 0.87 (0.69-1.08) | 0.10 |
| Palm or sole lesions | 8 (40%) | 74 (33%) | 1.19 (0.68-2.11) | 0.54 |
| Anal lesions | 5 (25%) | 30 (13%) | 1.81 (0.79-4.14) | 0.19 |
| Genital lesions | 16 (80%) | 91 (41%) | 1.95 (1.49-2.55) | 0.001 |
| Oral lesions | 8 (40%) | 49 (22%) | 1.79 (0.99-3.24) | 0.09 |
| Cough | 5 (25%) | 83 (37%) | 0.67 (0.31-1.45) | 0.29 |
| Sore throat | 12 (60%) | 94 (42%) | 1.41 (0.96-2.09) | 0.13 |
| Vomiting/nausea | 3 (15%) | 43 (19%) | 0.76 (0.26-2.24) | 0.68 |
| Asthenia | 12 (60%) | 139 (62%) | 0.96 (0.66-1.39) | 0.83 |
| Myalgia | 11 (55%) | 77 (35%) | 1.58 (1.02-2.44) | 0.08 |
| Conjunctivitis | 1 (5%) | 6 (3%) | 1.6 (0.2-12.64) | 0.56 |
| Eye itching | 4 (20%) | 37 (17%) | 1.18 (0.47-2.97) | 0.68 |
| Chills/sweats | 10 (50%) | 141 (63%) | 0.79 (0.5-1.24) | 0.26 |
| Headache | 11 (55%) | 104 (47%) | 1.17 (0.77-1.79) | 0.48 |
| Light sensitivity | 4 (20%) | 46 (21%) | 0.95 (0.38-2.38) | 0.9 |
| Confined to bed | 2 (10%) | 17 (8%) | 1.24 (0.31-5.01) | 0.67 |
| Diarrhea | 2 (10%) | 33 (15%) | 0.66 (0.17-2.55) | 0.61 |
| Rectal pain/bleeding | 1 (5%) | 7 (3%) | 1.4 (0.18-10.82) | 0.64 |
